# Evolutionary dynamics at the tumor edge reveals metabolic imaging biomarkers

**DOI:** 10.1101/2020.10.06.20204461

**Authors:** Juan Jiménez-Sánchez, Jesús J. Bosque, Germán A. Jiménez Londoño, David Molina-García, Álvaro Martínez, Julián Pérez-Beteta, Carmen Ortega-Sabater, Antonio F. Honguero Martínez, Ana M. García Vicente, Gabriel F. Calvo, Víctor M. Pérez-García

**Affiliations:** Mathematical Oncology Laboratory, Universidad de Castilla-La Mancha, Spain; Department of Mathematics, Universidad de Cádiz, Spain; Thoracic Surgery Unit, Hospital General Universitario de Albacete, Spain; Nuclear Medicine Unit, Hospital General Universitario de Ciudad Real, Spain

**Author notes:** G.F.C. and V.M.P.-G. proposed the hypothesis and designed the research; J.P.-B., V.M.P.-G., A.F.H.M., G.A.J.L. and A.M.G.-V. collected and processed the data and analyzed the medical implications; J.J.-S., J.J.B., G.F.C, D.M.-G. and A.M. performed the research and analyzed the data; J.J.-S., J.J.B., G.F.C. and V.M.P-G. drafted the paper. All authors read and approved the manuscript. J.J.-S. and J.J.B. contributed equally to this work. V.M.P.-G. and G.F.C. were both co-senior authors of this work.

**Keywords:** cancer, ^18^F-FDG PET/CT, evolutionary dynamics, prognostic biomarker

## Abstract

Human cancers are biologically and morphologically heterogeneous. A variety of clonal populations emerge within these neoplasms and their interaction leads to complex spatio-temporal dynamics during tumor growth. We studied the reshaping of metabolic activity in human cancers by means of continuous and discrete mathematical models, and matched the results to positron emission tomography (PET) imaging data. Our models revealed that the location of increasingly active proliferative cellular spots progressively drifted from the center of the tumor to the periphery, as a result of the competition between gradually more aggressive phenotypes. This computational finding led to the development of a metric, the NPAC, based on the distance from the location of peak activity (proliferation) to the tumor centroid. The NPAC metric can be computed for human patients using ^18^F-FDG PET/CT images where the voxel of maximum uptake (SUV_max_) is taken as the point of peak activity. Two datasets of ^18^F-FDG PET/CT images were collected, one from 61 breast cancer patients and another from 161 non-small-cell lung cancer patients. In both cohorts, survival analyses were carried out for the NPAC and for other classical PET/CT-based biomarkers, finding that the former had a high prognostic value, outperforming the latter. In summary, our work offers new insights into the evolutionary mechanisms behind tumor progression and provides a PET/CT-based biomarker with clinical applicability.

**Significance Statement:** Through the use of different *in silico* modeling approaches capturing tumor heterogeneity, we predicted that areas of high metabolic activity would shift towards the periphery as tumors become more malignant. To confirm the prediction and provide clinical value for the finding, we took ^18^F-FDG PET images of breast cancers and non-small-cell lung cancers, where we measured the distance from the point of maximum activity to the tumor centroid, and normalized it by a surrogate of the volume. We show that this metric has a high prognostic value for both malignancies and outperforms other classical PET-based metabolic biomarkers used in oncology.

---

Human cancers are genetically and morphologically heterogeneous (1, 2). This is generally attributed to the evolutionary dynamics of different clonal cell populations co-existing in the tumor ecosystem and undergoing stochastic branching processes over time (3–5). Successively acquired driver mutations, somatic alterations, and non-genetic modifications may confer increased fitness on certain cancer cell phenotypes, which subsequently outcompete those that do not experience such selection benefits within their microenvironment (4, 6, 7). Cells with specific advantageous traits may not show uniform spatial distribution across the tumor, particularly in large tumors. In fact, trade-offs exist that preclude the occurrence of optimal phenotypes, as exemplified by the hallmarks of cancer (8), and thus only local selection is expected to take place. This produces the spatial phenotypic diversity found in primary tumors and distant metastases (9).

Sustained metabolic reorganization during tumor progression, due to bioenergetically very demanding processes such as rapid proliferation, is a major hallmark of cancer (8, 10). This gives rise to a global metabolic plasticity and fitness optimization that confers evolutionary advantages under specific selective pressures, such as hypoxia (11). Positron emission tomography (PET) has been proposed as a way to assess macroscopic tumor heterogeneity in human patients (12). The technique is used in clinical practice with the radiotracer ^18^F-Fluorodeoxyglucose (^18^F-FDG) (13), which is an analog of glucose, and thus a marker of glycolysis (14). The altered tumor metabolism leads to an upregulation of glycolysis and an increase in glucose consumption (15). This happens even in the presence of oxygen and is referred to as the Warburg effect. Even though this process is energetically inefficient (16), cancer cells may find it beneficial to satisfy the biomass demands required by their high proliferation rates (17). This is confirmed by studies that relate the uptake of ^18^F-FDG in PET images to proliferation markers (18, 19). Therefore, the spatial map of glucose consumption provided by ^18^F-FDG PET images, as measured at each voxel by the standardized uptake value (SUV), is of great utility in portraying the spatial distribution of proliferation within the tumor.

The degree and impact of intertumor diversity and intratumor heterogeneity in patients has driven the need for quantitative frameworks to account for this variability (20). We considered how the metabolic activity might be distributed inside the tumor and how that information could be related to ^18^F-FDG PET images. Specifically, we looked at whether the location of prominent proliferation spots, as measured by the voxel of maximum radiotracer uptake (SUV_max_), could convey information about patient prognosis. We did this by analyzing how these spots changed over time and space within the tumor *in silico* using two mathematical models of different levels of complexity. The implications of these results, summarized in the definition of a novel prognostic biomarker, were tested on datasets of breast and lung cancer patients.

## Results

### Phenotype variability supports a drift of the peak metabolic activity towards the tumor boundary

To describe the emergence of metabolic heterogeneity, we studied *in silico* a simple biological scenario assuming the tumor to be composed of a clonal population of cells that can migrate, proliferate until the physical space is full, and die. To account for phenotypic heterogeneity, a transition probability that a cell proliferating at a rate *ρ* could increase or decrease its rate was introduced. The mathematical model used was a continuous non-local Fisher-Kolmogorov-type equation (21) which considered tumor cell population to be structured both by a spatial position vector **x** ∈ Ω ⊂ ℝ^3^, inside a domain Ω, and a proliferation rate *ρ* ∈ [0, *ρ*_m_], where *ρ*_m_ is a maximum proliferation rate. Let *u* = *u*(**x**, *ρ, t*) denote the cell density function, such that *u*(**x**, *ρ, t*) *d*^3^**x** *dρ* represents the number of tumor cells that, at time *t*, have a proliferation rate *ρ* at point **x**. We modeled the dynamics of *u*(**x**, *ρ, t*) via the following migration-proliferation integro-differential equation

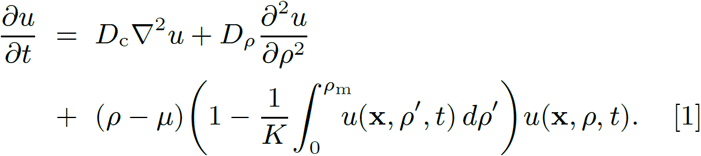

The first term accounts for cell migration with a diffusion constant *D*_c_ > 0. The second term captures the effect of non-genetic instability, mediated by fluctuations in the proliferation phenotype occurring with a diffusion constant *D*_*ρ*_ > 0. Note that the proliferation phenotype is a hallmark in tumors resulting from alterations in growth regulation (8). The third term comprises two main factors. The first includes the proliferation rate *ρ* minus a constant death rate *µ* > 0; those cells having a larger factor *ρ* − *µ* will tend to display a fitness advantage unless exogenous mechanisms (e.g. cytotoxic drugs targeting actively dividing cells) exert a negative selection effect on the phenotype. The second factor consists of a non-local logistic form with a carrying capacity *K* > 0. This factor represents the interplay between intratumor subpopulations with different proliferations competing for the available space.

A number of quantities are useful for summarizing the information contained in Eq. [1]. The first is the marginal cell density 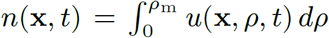, with typical radially symmetric profiles as shown in Fig. 1*A*. The second is the proliferation density ℳ (**x**, *t*) (see Eq. [2] in ‘Materials and Methods’), which gives the spatio-temporal proliferation map and allows the tumor regions with high metabolic activity to be identified. Figure 1*B* depicts ℳ (**x**, *t*)*/K* and shows how peak activity shifts from the tumor centroid towards the boundary as it grows *in silico*. This observed displacement, which was found to be linear with time, was quantified using two metrics. The first was the distance from the activity peak, corresponding to the point of maximum proliferation, to the tumor centroid. We named this metric PAC. The second was the normalized PAC (NPAC), defined as the ratio between PAC and the mean metabolic radius of the tumor, *R*_met_ (see Fig. 1*C*), which is thus size-independent. Simulations of Eqs. [1] showed that, during the early stages of the natural history of the tumor, the metric PAC was found to be zero or very small. However, as the inner regions were filled with cells, PAC increased linearly with time (see Fig. 1*D*). NPAC changed steadily from zero to one, since the maximum proliferation spot can only occur between the tumor center and its edge, indicating that this peak will move from the central regions of the tumor to its boundaries (see inset in Fig. 1*D*).

**Fig. 1.**
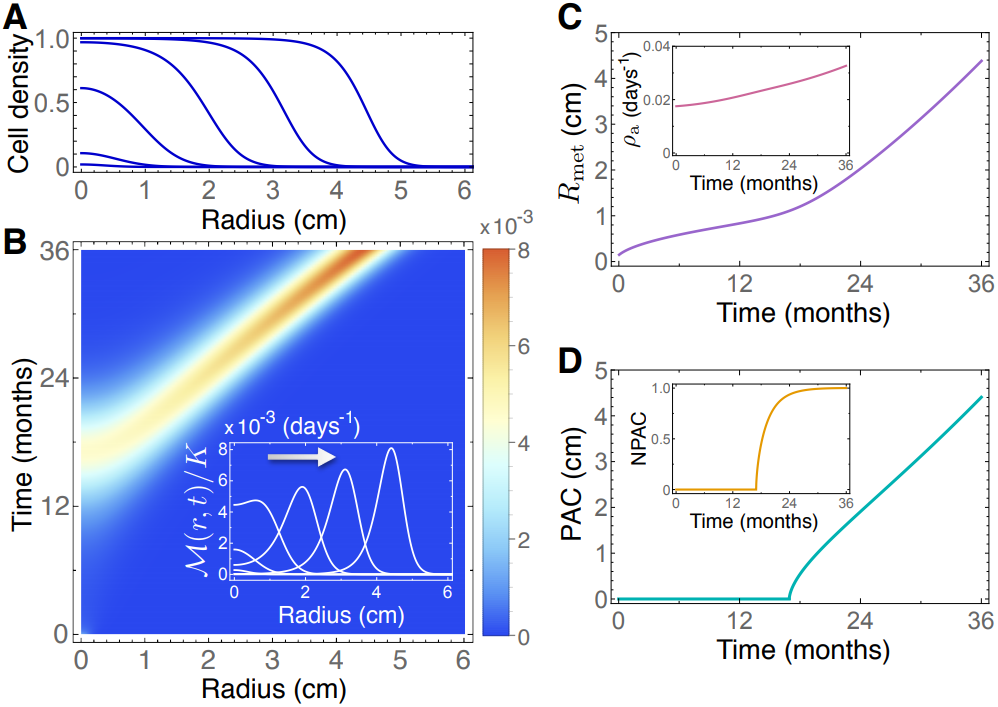
Non-local Fisher-Kolmogorov model [1] predicts a drift of the peak metabolic activity from the tumor centroid to the periphery with time. (*A*) Normalized cell density *n*(**x**, *t*_*j*_) */K* at *t*_*j*_ = 6, 12, 18, 24, 30 and 36 months (from left to right) for a radially symmetric tumor. (*B*) Pseudocolor plots of the normalized spatio-temporal proliferation density ℳ (**x**, *t*) and profiles (inset) of ℳ (**x**, *t*_*j*_) calculated at *t*_*j*_ = 6, 12, 18, 24, 30 and 36 months. (*C*) Mean metabolic radius *R*_met_(*t*) and (inset) average proliferation rate *ρ*_a_(*t*). (*D*) Variation over time of the distance from the tumor centroid to the point of maximum proliferation (PAC) and (inset) normalized PAC by the mean metabolic radius (NPAC). Simulation parameters are listed in ‘Materials and Methods’.

The simulations of Eqs. [1] revealed other noteworthy effects. Firstly, the amplitude of the maximum peak in ℳ (**x**, *t*) grew with time, meaning that tumors at later stages of their evolutionary history have larger peak activity values. This was in line with the frequently observed association between SUV_max_ and prognosis observed for different tumor histologies (22, 23). Secondly, the distribution of the proliferation rates displayed sustained growth towards higher values of *ρ*, reflected in the average proliferation rate *ρ*_a_ (see the inset of Fig. 1*C*). The growth of the tumor proliferation rate with time (size) has been experimentally observed in other studies (24).

### Genotype evolutionary dynamics supports the drift of tumor peak metabolic activity towards the boundary

We next resorted to a more complex and realistic biological scenario accounting for genotypic alterations. We did this by considering a stochastic discrete model based on fundamental cell features. At the cellular level, cancer cells can be characterized by four deregulated processes: proliferation, migration, mutation and death. These processes can be easily implemented as rules in a discrete mathematical model to mimic the main characteristics of the real system, with the drawback of facing high computational cost, especially when simulating clinically relevant volumes (25). To overcome this problem, we developed a hybrid stochastic mesoscale model of tumor growth that allowed clinically-relevant tumor sizes to be simulated while retaining the basic cancer hallmarks (24).

The model was parametrized for two of the most prominent cancer types, namely breast and lung cancer (non-small-cell lung carcinoma, NSCLC). A summary of our available data can be seen in Table 1 in ‘Materials and Methods’. Mutational landscapes were constructed based on a simplification of their known mutational spectra. Alterations in EGFR and ALK, which are strongly associated with non-squamous lung adenocarcinoma, were considered to model NSCLC, while driver mutations in PIK3CA and TP53 were considered for breast cancer (26–29). Therefore, the mutational tree in both types of tumors simulated had two possible altered genes, leading to four possible combinations or ‘genotypes’ that define four different clonal populations. Basal rates associated a characteristic time to each basic cell process, and mutation weights determined how these basal rates were affected once a given alteration was acquired. Mutation weights were taken to contribute equally for all alterations, and their effect was cumulative, so that a cell carrying two alterations simultaneously would perform basic processes with a double advantage. Thus, the stochastic mesoscopic model provided a richer scenario to explore intratumoral heterogeneity during tumor growth.

**Table 1.** Stochastic model parameters.

| Parameter | Meaning | Value | Reference |
| --- | --- | --- | --- |
| $L$ | Number of voxels per side | 80 | - |
| $\Delta x$ | Voxel side length [mm] | 1 | (52) |
| $\Delta t$ | Time step length [hours] | 24 | - |
| $K$ | Carrying capacity per voxel [cells] | $2 \cdot 10^5$ | (53) |
| $N_t$ | Threshold cell number | $0.2 \cdot K$ | - |
| $N_0$ | Initial population [cells] | 1 | - |
| $V_{\text{end}}$ | Maximum reachable tumor volume [cm<sup>3</sup>] | 50 (breast)<br>120 (lung) | (54) |
| $V_{\text{diag}}$ | Tumor volume at diagnosis [cm<sup>3</sup>] | 0.3-5 (breast)<br>0.2-15 (lung) | (55)<br>(56) |

We ran 100 simulations of breast cancer and 100 of NSCLC with random parameters uniformly sampled from the ranges in Tables 1 and 2 (see ‘Materials and Methods’). Cell number, activity (number of newborn cells) and most abundant clonal population were calculated for each voxel and time step (Fig. 2). Tumor volumes were measured from the number of voxels containing more than a threshold number of cells *N*_*t*_ (Table 1), and the mean spherical radii (MSR) computed from these.

**Table 2.**
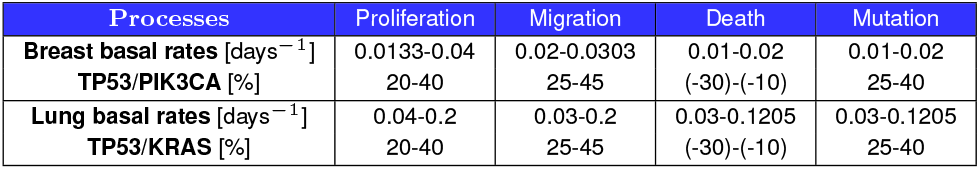
Basal rates and mutation weights.

**Fig. 2.**
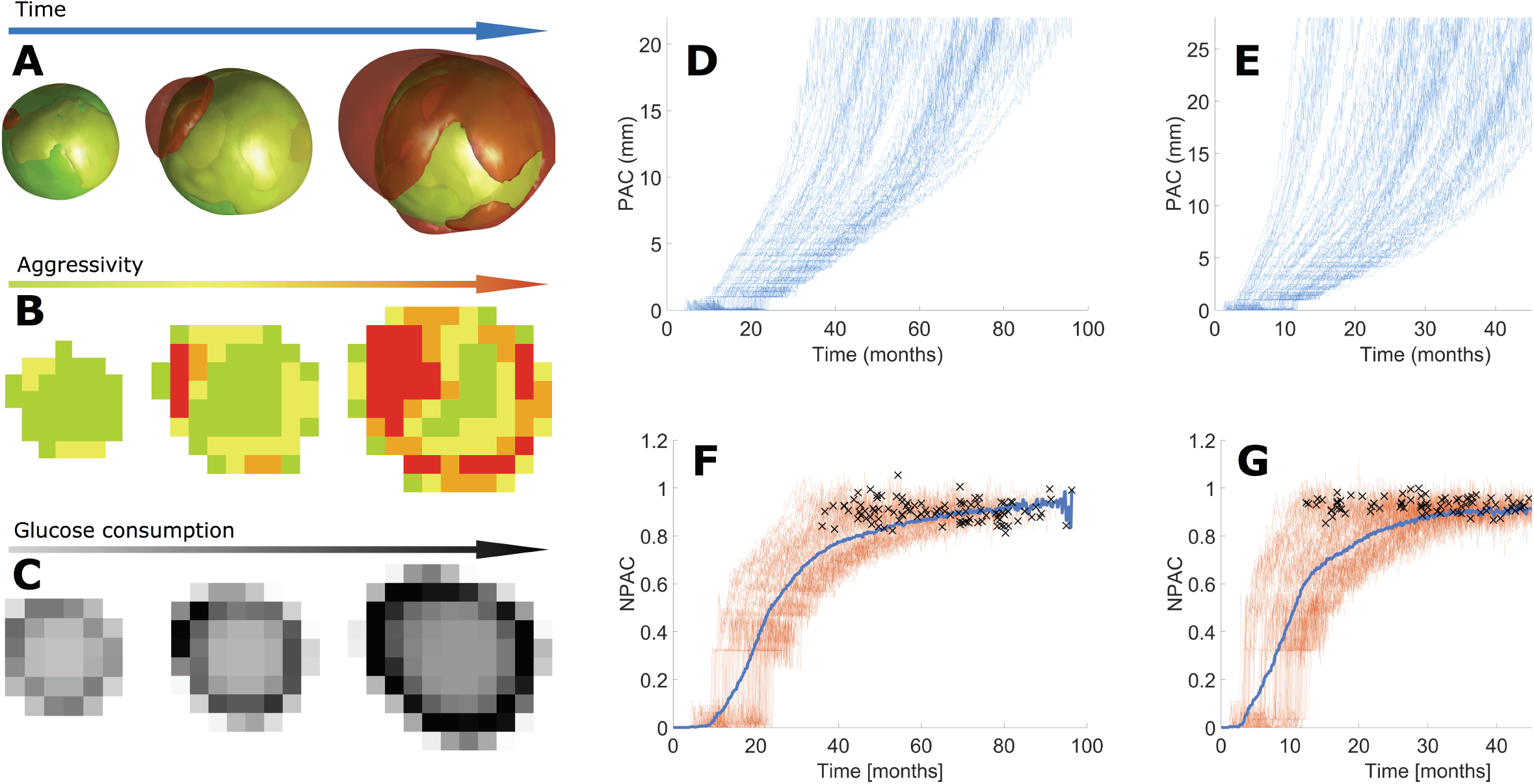
Hybrid stochastic mesoscale model shows that competition among progressively more aggressive phenotypes is pushed to the edge. (*A*) 3D volume renderings at different time frames (from left to right: 78, 85 and 92% of simulation) of a simulation of breast cancer growth depicting clonal populations within the tumor. Color of cell populations ranges from green (less aggressive) to red (more aggressive). Rates are: proliferation 0.0315 day^−1^, death 0.0157 day^−1^, mutation 0.0160 day^−1^, and migration 0.0235 day^−1^. (*B*) Central sections for the same simulation and time frames as in (*A*) showing the most abundant clonal population per voxel. (*C*) Central section of tumor activity for the same time frames as in (*A*). (*D, E*) PAC progression for every simulation of breast cancer (*D*) and NSCLC (*E*). (*F, G*) Longitudinal NPAC dynamics for simulations of breast cancer (*F*) and NSCLC (*G*) growth, with individual runs colored in reddish orange and all-simulation averaged NPAC in blue; dark crosses depict the time points at which each simulation ended.

As cells mutated *in silico*, new clonal populations emerged with higher, more advantageous migration and proliferation rates. These new clones increased their relative abundance in the tumor, eventually becoming fixed in the system. As the tumors grew larger, cell division occurred preferentially at the tumor periphery. This was as expected, since inner voxels became progressively filled with cells that prevented them from proliferating. Voxels where the most aggressive clonal population was more abundant were associated with spots of maximum proliferation. Therefore, *evolution was pushed towards the tumor edge*: cells with higher fitness (specially those having higher proliferation rates) appeared farther from the tumor center as they grew. At each time step, the maximum proliferation spot was identified as the voxel with the largest number of cell births, and its distance to the tumor centroid (PAC) calculated. Figures 2*D, E* show a monotonic increase of PAC with time for both histologies. Normalizing with respect to the MSR to get the NPAC showed that the point of maximum proliferation was displaced towards the boundary (Figs. 2*F, G*) in all the simulations performed. The only difference between simulations was the time that the maximum proliferation spot took to reach the edge. Thus, NPAC was predicted to be a robust property related to the evolutionary state of the disease.

### PET imaging data confirms evolutionary dynamics of the peak metabolic activity and validates related biomarkers

The computational results suggested that NPAC might contain meaningful prognostic information. In the clinical setting, the metabolic activity distribution of the tumor can be evaluated by means of F-FDG PET, which reflects the biological processes taking place at a lower level (30), and is frequently used on newly diagnosed breast cancer and NSCLC patients. To confirm or refute the theoretical predictions, we performed a study on our patient cohort (see ‘Materials and Methods’). For each patient, the tumor was delineated in the images and the locations of centroids and SUV_max_ were obtained from the segmented distribution. The metabolic tumor volume (MTV), total lesion glycolysis (TLG; integral of the SUV distribution over the volume), and NPAC metrics were calculated for all tumors for both histologies. Two typical examples of ^18^F-FDG PET images from breast cancer patients are shown in Fig. 3(A) and (B), respectively. Small values of NPAC, with SUV_max_ close to the tumor centroid as in Fig. 3(B,I), were expected to correspond to less developed disease, in accordance with the previous theoretical framework. In contrast, the cases shown in Fig. 3(C,J) with SUV_max_ displaced in relation to the centroid, would correspond to tumors with a poorer prognosis.

**Fig. 3.**
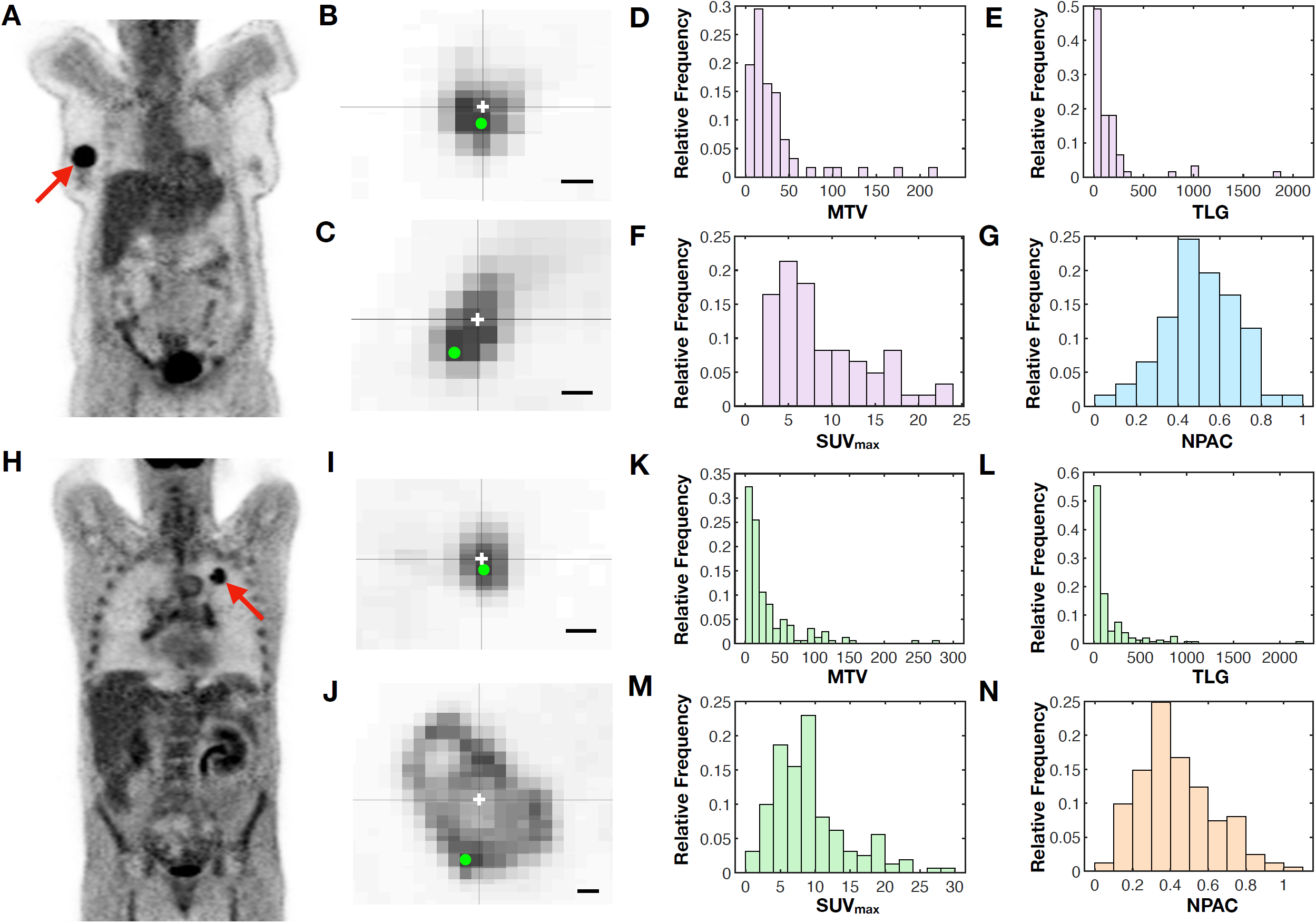
Analysis of NPAC in ^18^F-FDG PET images reveals well-behaved distributions with prognostic potential. (*A,H*) Examples of PET images for breast (*A*) and NSCLC (*H*) patients in our dataset. (*B,C,I,J*) Two-dimensional slices from patients with small (*B,I*) and large (*C,J*) NPAC values for breast (*B,C*) and NSCLC (*I,J*) patients. The centroid of each segmented lesion and the voxel of SUV_max_ are marked with a white cross and a green dot respectively. Scale bar lengths are 1 cm. (*D-G,K-N*) Histograms showing the distributions of metabolic tumor volume (*D,K*), total lesion glycolysis (*E,L*), SUV_max_ (*F,M*) and NPAC (*G,N*) for breast cancer (*D-G*) and NSCLC (*K-N*) patients in our datasets.

The histograms in Fig. 3(D-G, K-N) depict the distributions of MTV, TLG, SUV_max_ and NPAC for both histologies. It is noteworthy that the NPAC has a more regular distribution than the other PET-based measures, with definite values between 0 and 1 and a centered mean (breast cancer: 0.51 ± 0.18, median 0.50; NSCLC: 0.43 ± 0.2, median 0.39). It is clear from Fig. 3(G,N) that at the time of diagnosis the point of maximum uptake is typically located away from the geometrical center of the tumor.

The classical measures (MTV, TLG, SUV_max_) are known to be prognostic biomarkers in breast cancer and NSCLC (22, 23). For these variables we performed Kaplan-Meier analyses on overall survival (OS) and disease-free survival (DFS) (Fig. S5). All of the variables had prognostic value in the breast cancer cohort, but only MTV returned significant results (p-value<0.05) in the NSCLC cohort.

We then tested the prognostic value of NPAC by Kaplan-Meier analyses with OS and DFS as endpoints (see ‘Materials and Methods’). Results for the best splitting thresholds are shown in Fig. 4. For the breast cancer cohort, NPAC showed robust results in terms of OS, with a best splitting threshold in both OS and DFS of NPAC=0.499 (Fig. 4A,C). Interestingly for OS, the most relevant metric, the C-index reached an outstanding value of 1 (for DFS it was 0.899). Thus, no patients with tumors having their SUV_max_ closer than half the radius (*n*=30) died from the disease. In NSCLC, NPAC separated the patients well, and the best splitting threshold, NPAC=0.64, led to a C-index of 0.875 for OS. The separation in median OS between groups was 57.33 months, while in DFS it was 36.62 months.

**Fig. 4.**
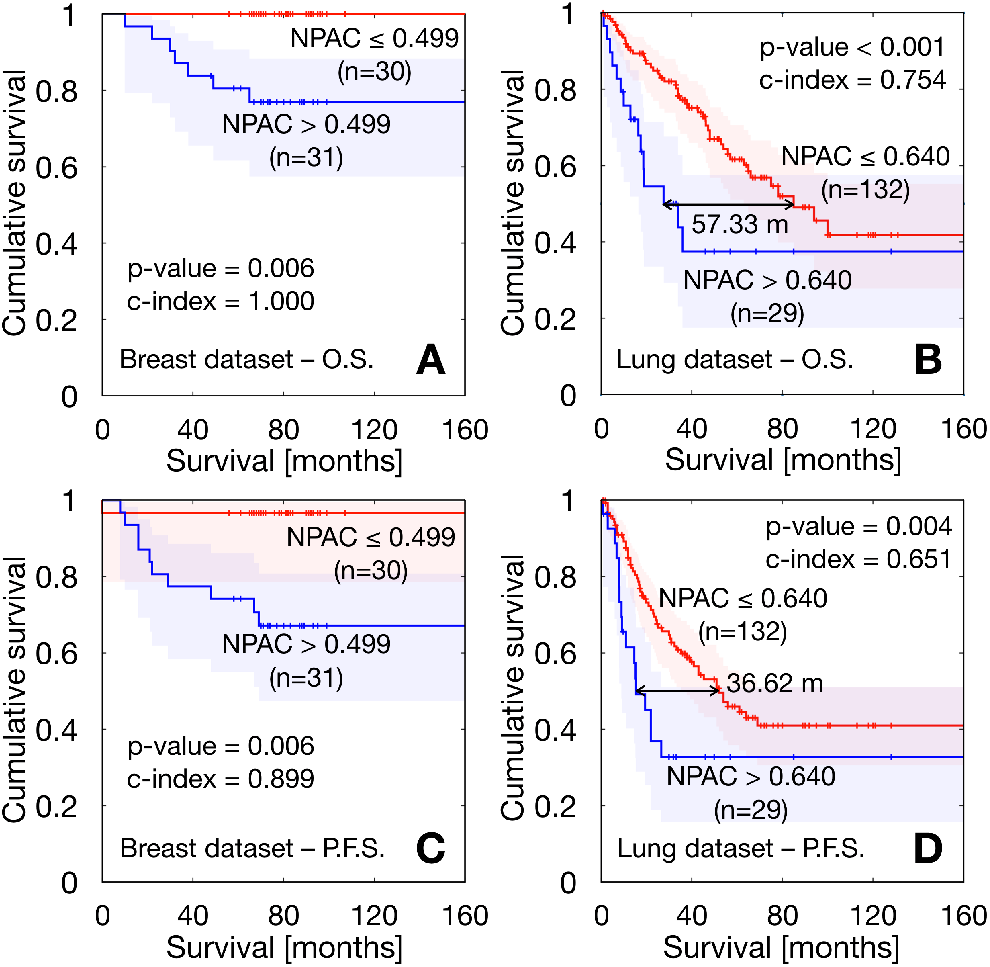
Kaplan-Meier curves obtained for best splitting thresholds corresponding to the NPAC metric. (*A*) Overall survival in breast cancer cohort. (*B*) Overall survival in NSCLC cohort. (*C*) Disease-free survival in breast cancer cohort. (*D*) Disease-free survival in NSCLC cohort.

These results show the strength of the NPAC as a prognostic biomarker in comparison with the classical metrics. For OS in the breast cancer cohort, only MTV approached the performance of NPAC; however, the C-index for NPAC (C-index = 1) outperformed the result for MTV (C-index = 0.875). For DFS, none of the classical variables showed non-isolated thresholds leading to a statistically significant association between subgroups. In the lung cancer cohort, the NPAC metric outperformed, once again, the prognostic value of the classical variables. Regarding OS, there were ranges of thresholds of MTV, TLG and SUV_max_ leading to statistically significant results with best C-indexes of 0.736 (MTV), 0.682 (TLG), 0.658 (SUV_max_) still substantially lower than the value obtained for NPAC (0.875). Results for DFS were again similar, with only MTV and TLG achieving significance, with best values of 0.638 (MTV) and 0.607 (TLG), but still underperforming NPAC, with a C-index of 0.651.

The results for correlations between the metabolic variables are shown in Fig. S7. NPAC was found to be independent of the classical metabolic variables.

## Discussion and conclusion

Heterogeneity is one of the hallmarks of tumor malignancy (1, 2). Many mathematical models have been constructed accounting for different aspects of the development of heterogeneity through evolutionary dynamical processes in a number of cancer types (31–33). We did not intend here to develop a universal mathematical model to describe every aspect of tumor growth progression, but rather to focus on understanding the basic dynamics of the peak of metabolic activity, due to the potential applicability of the results. Different levels of complexity were considered in each of the two complementary models constructed, and both led to the same conclusions: (i) tumors would evolve towards higher proliferation rate values, and (ii) peak metabolic activity would move towards the tumor edge as the tumor evolves with time.

It is interesting to note that conclusion (ii) (but not (i)) would have been arrived at through the use of a ‘classical’ local Fisher-Kolmogorov model (34). In the context of that mathematical model, proliferation is inhibited in areas of high cell density and higher proliferation areas would switch from the tumor core to the periphery, as is observed in the context of the non-local model Eq. (1). This is, therefore, a robust finding of the study, and it is likely to occur in other mathematical approaches. Conclusion (i) does not come as a surprise, since the maximum metabolic activity obtained from PET images (e.g. as measured by the SUV_max_), has been known to contain prognostic information in different cancer types (22, 23). Thus, the fact that SUV_max_ has a prognostic value in the clinical setting is compatible with the results of our models, where the activity grows with time over the tumor’s natural lifetime until a maximum value is reached.

Many studies have correlated either classical PET-derived metrics such as MTV, TLG (35, 36) or complex spatial features of the distribution of SUV values (37) with the outcome of the disease. However, no study has analyzed the prognostic value of metrics derived from the location of the peak of metabolic activity. The fact that this simple biomarker has a high prognostic value is remarkable and probably related to the robustness of the biological assumptions behind the mathematical models used to substantiate it. In fact, it is natural to expect that the presence of more aggressive glucose-avid cells, that might be unable to progress when located in saturated areas near the center of the tumor, may be a risk factor when placed in regions with much more capacity to settle and invade.

In our study we chose to take the voxel bearing the SUV_max_ as the location of maximum metabolic activity to compute NPAC. We could also have used SUV_peak_ (the maximum SUV appearing in the distribution when all the voxels are averaged with their 26 neighbors), which is thought to be more stable and to better define an extended region of high uptake (38). However, SUV_max_ is often placed in the area defined by SUV_peak_, thus leading to equivalent metrics (39). SUV_max_ is also easier to identify visually and is therefore easier to use in clinical practice, besides being the simplest option.

The fact that NPAC provides an accessible and powerful prognostic metric could be extended in different ways. Firstly, it would be valuable to look at whether changes in this biomarker might provide a robust indication of an increase in malignancy for initially indolent tumors (e.g. benign lung nodules, low-grade gliomas, etc.) undergoing a malignant transformation. Secondly, an intriguing open question would be to determine whether the rate at which NPAC changes, during patient follow-up, correlates with the occurrence and fixation of specific mutations. Finally, it may be the case that changes with time of this metric, after different treatment modalities, could help in assessing the response through sequential PET studies as a measure of how much NPAC is reduced.

Mathematical and computational models are progressively gaining their place among the tools that are used to study cancer (40). *In silico* models based on evolutionary dynamics may capture relevant aspects of tumor growth, and have been proven helpful in understanding tumor clonal heterogeneity, one of the main hallmarks of cancer (41). Mechanistic mathematical models of different levels of complexity have been shown to provide biomarkers of clinical significance (see e.g. (24, 42–49). This type of approach provides a rational alternative to radiomic and deep-learning studies, where a mechanistic explanation is often missing. The study described in this paper falls into the former category, demonstrating that an informed understanding of the system’s emergent properties can shed light on the deeper roots of its working.

It is worth mentioning that our mathematical approach, beyond its fundamental interest, has led to the proposal of a simple metric with high prognostic value that can be obtained from ^18^F-FDG PET studies. The NPAC biomarker was able to separate patients of breast cancer and non-small-cell lung cancer into two groups with significantly different survival (both overall and disease-free) and was proven to be more powerful than traditional ^18^F-FDG PET/CT biomarkers (MTV, TLG, SUV_max_) currently used in clinics. This demonstrated that the geometric location of the peak metabolic activity, and not only its value, contains information of clinical significance.

This study opens many new avenues for research. Firstly, the search for other biomarker definitions accounting for the location of peak metabolic activity. Secondly, it would be interesting to test our findings in other tumor histologies. PET is a mainstream technique, increasingly employed in clinics and in many imaging studies for which a broad spectrum of tumor histologies is available. Thus, the applicability of NPAC to other cancer types would be an interesting extension of our work.

In conclusion, by using two mathematical models incorporating evolutionary dynamics, we have shown that peak metabolic activity is expected to increase in magnitude and to move towards the tumor boundary as human solid tumors progress. On the basis of the theoretical predictions we have defined a metric, the NPAC, representing the normalized distance from the peak of activity to the tumor centroid, and validated it as a prognostic biomarker in lung and breast cancer patients using PET imaging datasets. The new biomarker outperformed classical PET-based biomarkers such as TLG, MTV and SUV_max_ and provides a notable example of mathematically-grounded research with applicability in oncology.

## Materials and Methods

### Patients

Our study was based on data from two different studies. Breast cancer patients were participants of a multicenter prospective study approved by the Institutional Review Board (IRB). Written informed consent was obtained from all patients. The inclusion criteria were: (1) newly-diagnosed locally-advanced breast cancer with clinical indication of neoadjuvant chemotherapy, (2) lesion uptake higher than background, (3) absence of distant metastases confirmed by other methods prior to the request of PET/CT for staging, and (4) breast lesion size of at least 2 cm. 61 patients (18% lobular carcinoma, 82% ductal carcinomas, 100% women, age rank 25-80, median 50 years) were included in this dataset. The TNM data were: 54% T2, 18% T3, 28% T4; 28% N0, 55% N1, 6% N2, 11% N3; 100% M0.

175 patients (153 men, 22 women, age rank 41-84, median 65 years) were included in the study from a dataset of lung cancer patients who received surgery in the period June 2007 to December 2016. Histologies were 63 squamous-cell carcinomas and 112 adenocarcinomas. Staging information was: 69 stage I, 70 stage II, 33 stage III, 3 stage IV. The N staging was 107 patients N0, 46 N1 and 22 N2. All patients had M0. PET protocol and machine were as in subgroup 1. The inclusion criterion was established that minimal lesion size should be greater than 2.0 cm. From those initial patients, 14 were removed due to the unavailability of survival data.

The PET machine was a dedicated whole-body PET/CT scanner (Discovery SDTE-16s, GE Medical Systems) in 3D mode. Image acquisition began 60 minutes after intravenous administration of approximately 370 MBq (10 mCi) of ^18^F-FDG; the images obtained had a voxel size of 5.47 × 5.47 × 3.27 mm^3^, with no gap between slices, and a matrix size of 128 × 128. The inclusion criteria considered only newly diagnosed patients with availability of pre-treatment PET/CT examination and a lesion uptake higher than background, absence of distant metastases, and a lesion size of at least 2 cm.

### ^18^F-FDG PET image analysis and computation of the relevant metrics

PET images in DICOM format were loaded into MATLAB for the image analysis. In each image, the tumor was manually selected and subsequently delineated in 3D by an automatic algorithm. For all images, we evaluated the metabolic tumor volume (MTV; volume of the delineated tumor), the total lesion glycolysis (TLG; integral of the SUV distribution over the volume) and the SUV_max_ value. The centroids of segmented tumors and the distances from them to the position of the respective SUV_max_ (PAC) were obtained. Considering a sphere having the same volume as the MTV, the MSR was calculated to serve as a linear surrogate of volume. We then normalized the PAC of every tumor to the size-independent NPAC metric.

### Kaplan-Meier statistics

We performed Kaplan-Meier analyses over these two cohorts of patients, using the Log-Rank and Breslow tests to assess the significance of the results. These methods compare two populations separated in terms of one parameter and study their statistical differences in survival. Specifically, overall survival (OS) and disease free survival (DFS) Kaplan-Meier analyses were performed. A 2-tailed significance level with p-value lower than 0.05 was applied. The hazard ratio (HR) and its adjusted 95% confidence interval (CI) was also computed for each threshold using Cox proportional hazards regression analysis.

### Splitting thresholds

For each variable, we searched for every value splitting the sample into two different subgroups, satisfying the condition that none of them be more than 5 times larger than the other. We then tested each of them as splitting thresholds through Kaplan-Meier analyses, obtaining the significance results shown in Figs. S1-S4. The best splitting threshold was chosen as the non-isolated significant value giving the lowest p-value in both Log-Rank and Breslow tests, as described in Ref. (44).

### Harrell’s C-index

To assess the accuracy of prognostic models, Harrell’s concordance index score was also computed (50). This method compares the survival of two populations of patients (best prognosis versus worst prognosis) by studying all possible combinations of individuals belonging to different groups. Then, the percentage of right guesses is the reported result. Concordance indexes were computed using the non-censored sample and ranged from 0 to 1, with 1 indicating a perfect model (a purely random guess would give a concordance index of 0.5).

### Variable correlations

Spearman correlation coefficients were used to assess the dependencies between pairs of variables. We considered significant correlation coefficients over 0.7 or below −0.7 as strong (direct or inverse respectively) correlations between variables. In this way we were able to exclude possible confounding effects in our analysis.

### Statistical software

SPSS (v. 22.0.00), MATLAB (R2018b, The MathWorks, Inc., Natick, MA, USA) and R (3.6.3) software was used for all statistical analyses.

### Non-local Fisher-Kolmogorov model and simulations

The migration-proliferation integro-differential equation [1] in radial coordinates was solved numerically using the method of lines (51) combined with Newton-Cotes integration formulas to deal with the non-local term. In the simulations displayed in Fig. 1, the computational domain consisted of a radial variable *r* ∈ [0, *R*_max_], where *R*_max_ = 7 cm was the maximum radius, and the proliferation rate *ρ* ∈ [0, *ρ*_m_], where the maximum proliferation rate was *ρ*_m_ = 0.06 day^−1^. The number of nodes in the discretized *r*-*ρ* mesh was 350 × 180. Additional parameters were *D*_c_ = 3.5 · 10^−4^ cm^2^/day, *D*_*ρ*_ = 1.3 · 10^−8^ day^−3^, *µ* = 4 · 10^−3^ day^−1^ and *K* = 8 · 10^7^ cells/cm^3^. The initial condition consisted of a highly localized lesion with a radius of 1 mm containing 10^5^ tumor cells and having a mean proliferation rate *ρ*_0_ = 1.7 · 10^−2^ day^−1^ and standard deviation *σ*_0_ = 3 · 10^−3^ day^−1^.

The general expression for the proliferation density is

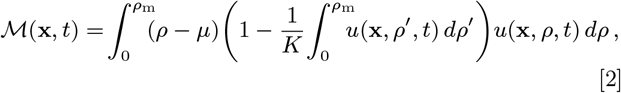

and was used to compute the plots shown in Fig. 1*B* assuming spherical symmetry.

In Fig. 1*C*, the mean metabolic radius was defined as

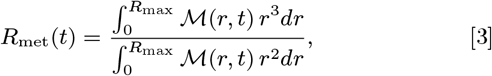

while the average proliferation rate was determined via

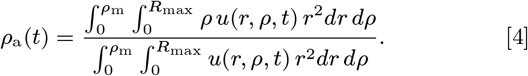

In Fig. 1*D*, the distance from the tumor centroid to the point of maximum proliferation (PAC) was calculated at each time step via expression [2]. The normalized PAC (NPAC) was computed by means of the ratio NPAC(*t*) = PAC(*t*)*/R*_met_(*t*).

## Data Availability

All data is included in the manuscript and supporting information.

