## Supporting Information for "Evolutionary dynamics at the tumor edge reveals metabolic imaging biomarkers"

PNAS
www.pnas.org

### Supplementary Information for

#### Evolutionary dynamics at the tumor edge reveals metabolic imaging biomarkers

**J. Jiménez-Sánchez, J.J. Bosque, G.A. Jiménez Londoño, D. Molina-García, A. Martínez, J. Pérez-Beteta, C. Ortega-Sabater, A.F. Honguero Martínez, A.M. García Vicente, G.F. Calvo and V.M. Pérez-García.**

##### This PDF file includes:

Supplementary text
Figs. S1 to S17
Legends for Movies S1 to S3
Legends for Dataset S1 to S2
SI References

##### Other supplementary materials for this manuscript include the following:

Movies S1 to S3
Datasets S1 to S2

#### Supporting Information Text

### 1. Stochastic model and Patient Survival Analysis

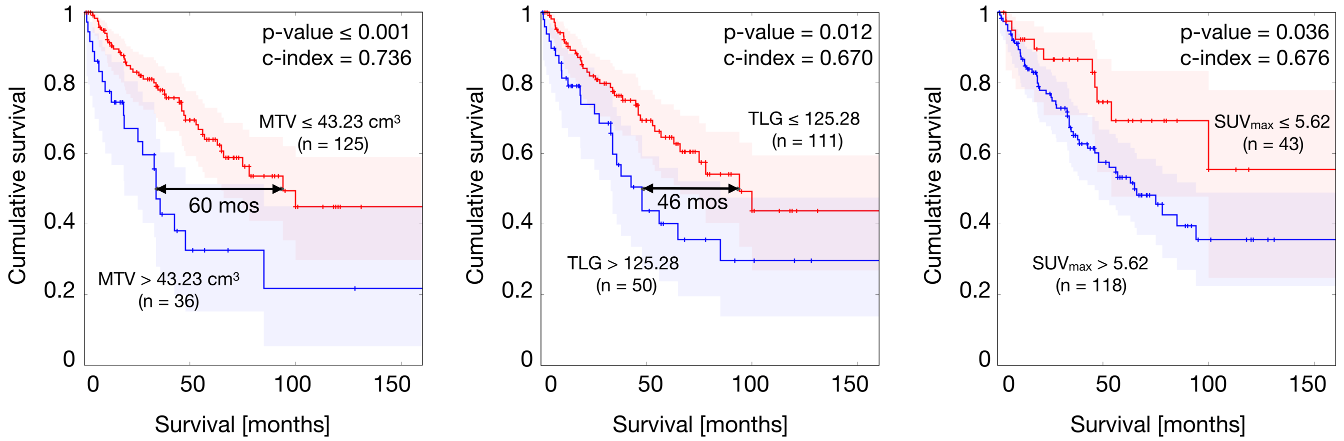


**Fig. S1.** Kaplan-Meier analyses for overall survival of the 161 NSCLC patients performed on metabolic tumor volume, total lesion glycolysis and $SUV_{max}$, respectively.

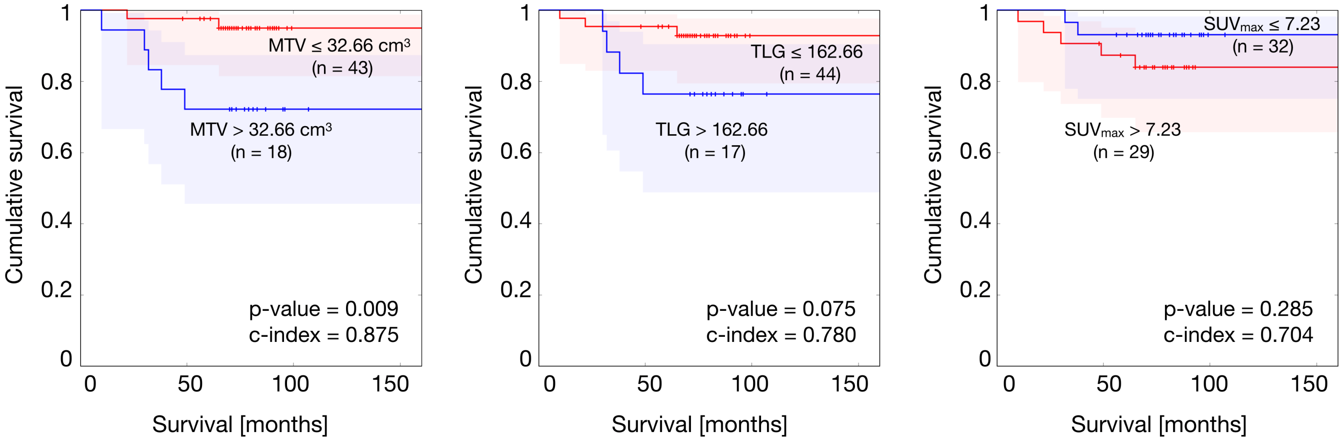


**Fig. S2.** Kaplan-Meier analyses for overall survival of the 61 breast patients performed on metabolic tumor volume, total lesion glycolysis and $SUV_{max}$, respectively.

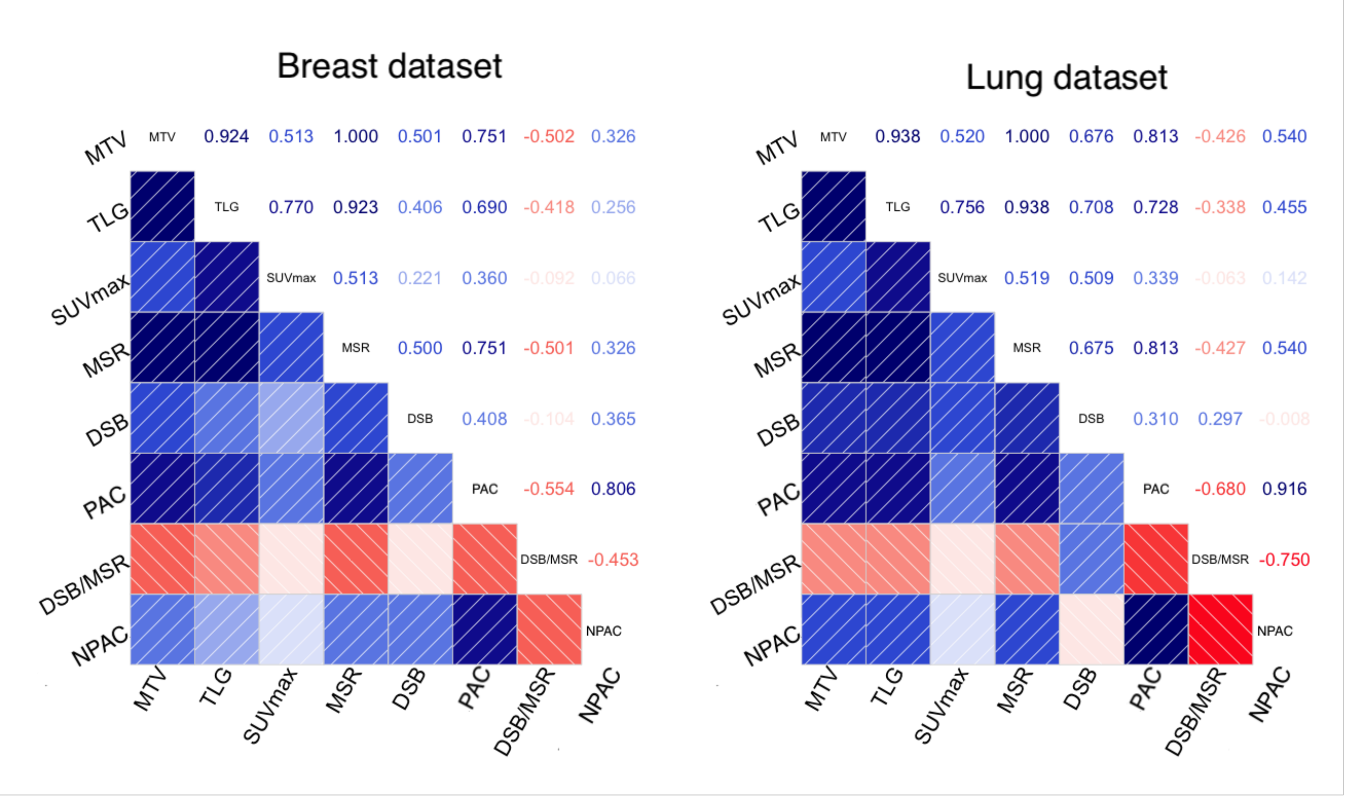


**Fig. S3.** Analyses of correlation of the different variables considered in the study for the two cohorts: Metabolic Tumor Volume (MTV), Total Lesion Glycolysis (TLG), Maximum Standardized Uptake Value ($SUV_{max}$), Mean Spherical Radius (MSR), Distance from $SUV_{max}$ to tumor Border (DSB), distance from $SUV_{max}$ to tumor centroid (PAC), DSB normalized by MSR (DSB/MSR), and PAC normalized by MSR (NPAC).

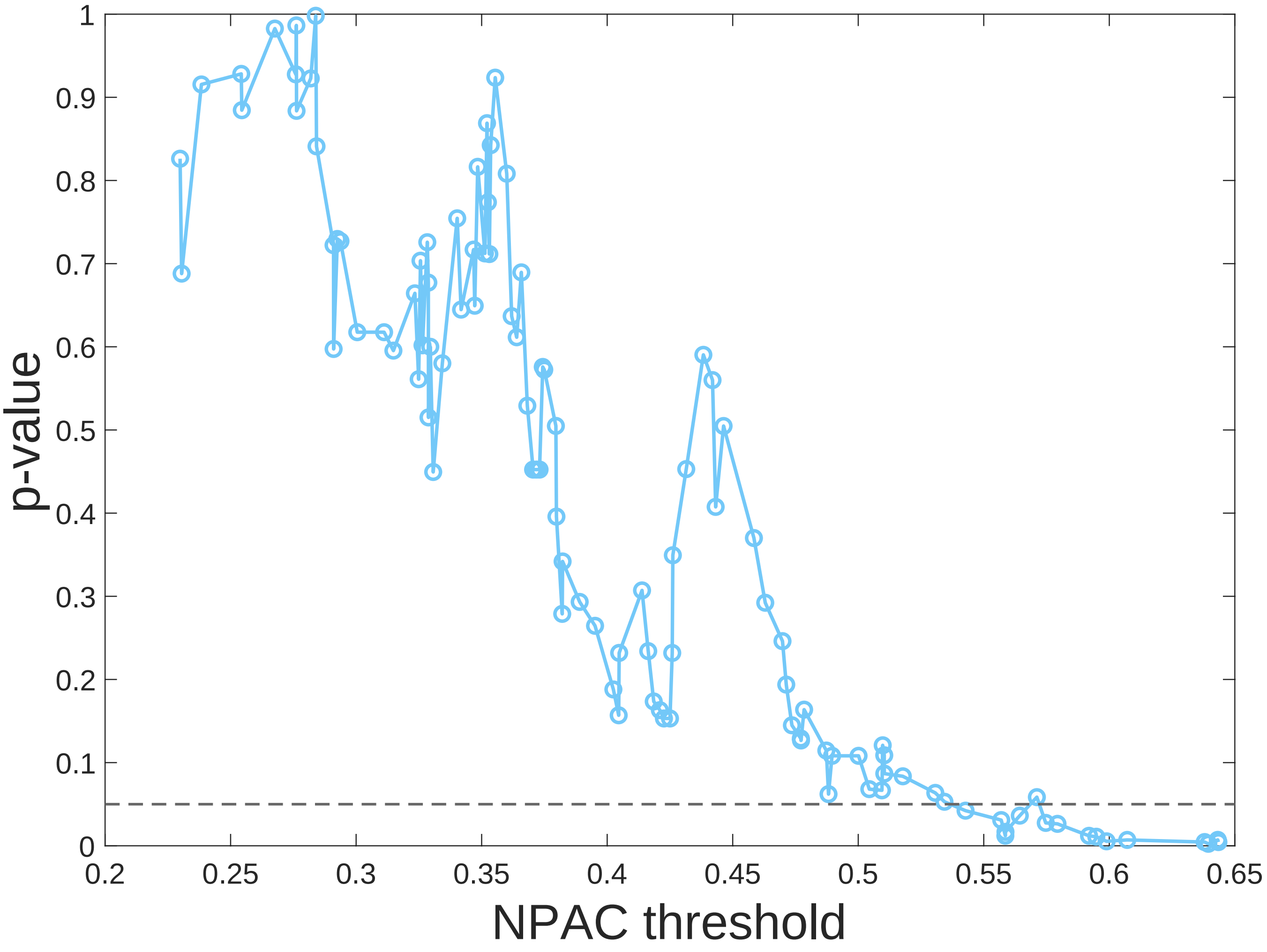


**Fig. S4.** To perform the survival analysis we evaluate all the possible thresholds of NPAC, separating the patients in two different groups according to their NPAC in relation to each threshold. For each of those thresholds, a Kaplan-Meier analysis is performed and the significance of the separation among the two curves is measured by their p-value. In the figure, the different p-values are plotted for the different NPAC thresholds evaluated for the NSCLC dataset. Only groups where the number of patients in the largest one is at most five times bigger than in the smallest one are considered. The level of statistical significance (p-value$\leq$0.05) is represented by the dashed line. The NPAC is able to discriminate two significantly different groups with disparate survivals in a robust manner.

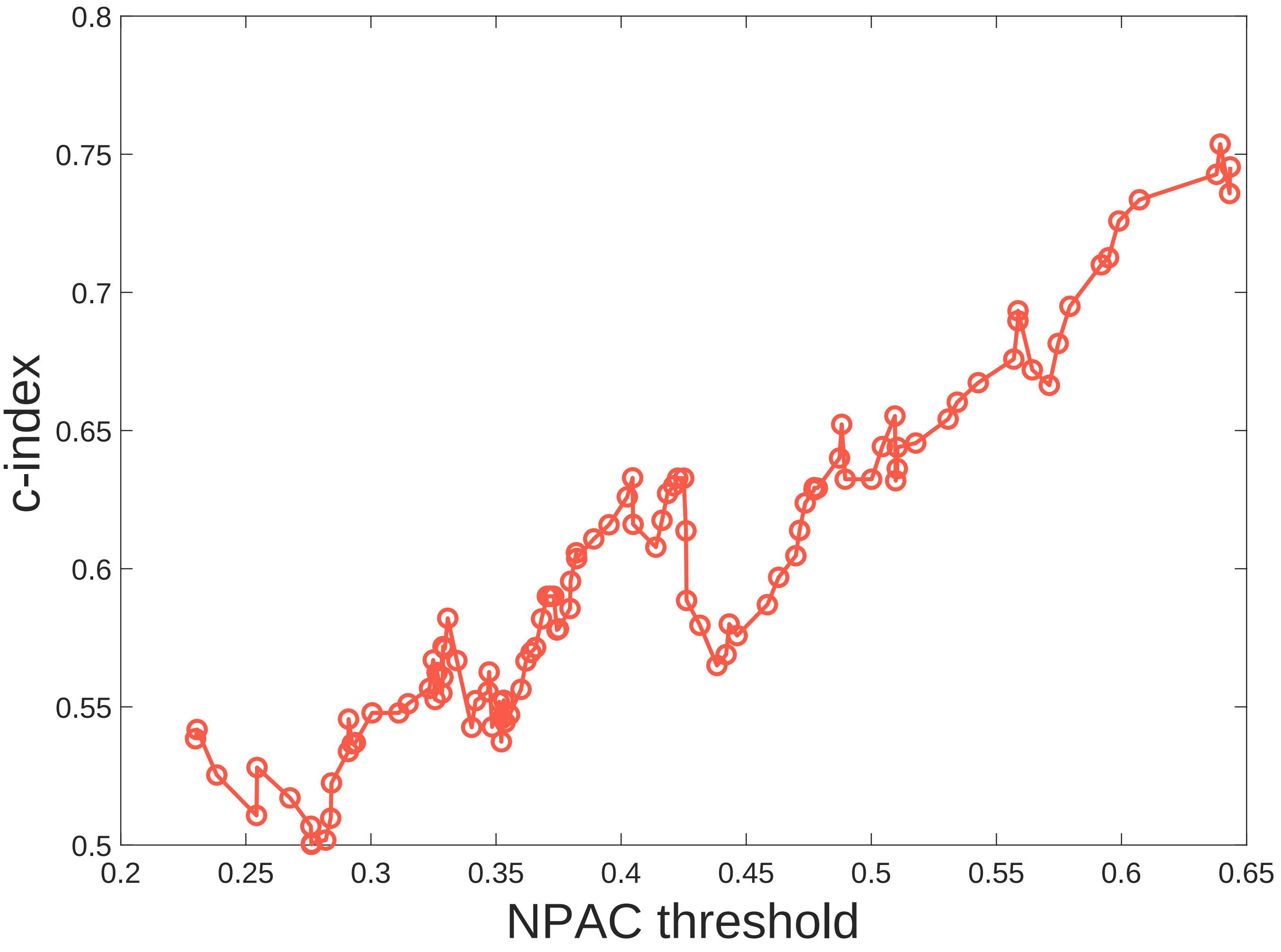


**Fig. S5.** To perform the survival analysis we evaluate all the possible thresholds of NPAC, separating the patients in two different groups according to their NPAC in relation to each threshold. For each of those thresholds, a Kaplan-Meier analysis is performed and the Harrell's c-index is calculated to evaluate the risk associated to patients with dissimilar NPAC. In the figure, the different c-indexes are plotted for the different NPAC thresholds evaluated for the NSCLC dataset. Only groups where the number of patients in the largest one is at most five times bigger than in the smallest one are considered. The separation between the two groups, given by high values of the NPAC, correctly discriminates the risks (disparate survival times) between both groups, as evidenced by the high values obtained for the c-indexes.

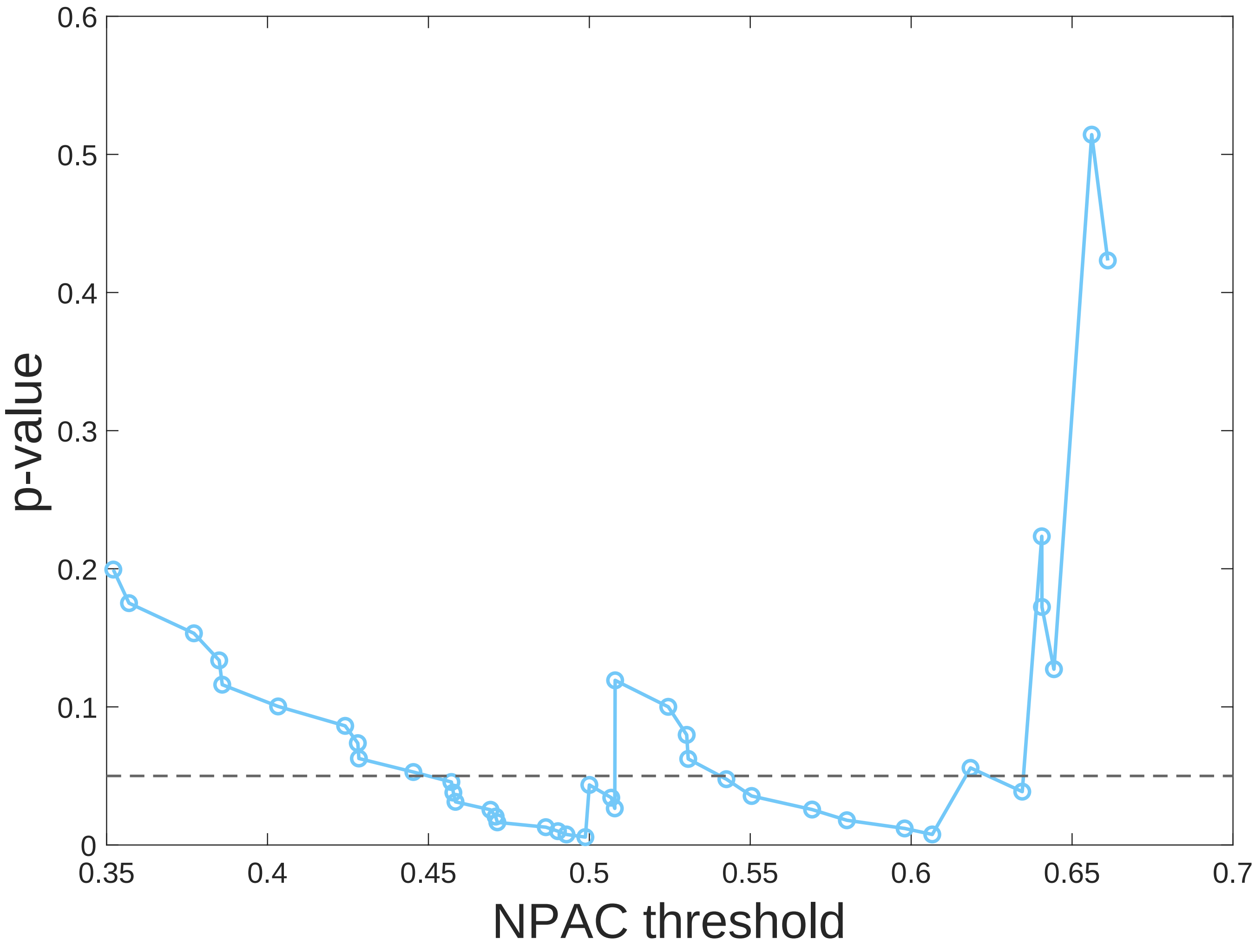


**Fig. S6.** To perform the survival analysis we evaluate all the possible thresholds of NPAC, separating the breast cancer patients in two different groups according to their NPAC in relation to each threshold. For each of those thresholds, a Kaplan-Meier analysis is performed and the significance of the separation among the two curves is measured by their p-value. In the figure, the different p-values are plotted for the different NPAC thresholds evaluated. Only groups where the number of patients in the largest one is at most five times bigger than in the smallest one are considered. The level of statistical significance (p-value$\leq$0.05) is represented by the dashed line. The NPAC is able to discriminate two significantly different groups with disparate survivals in a robust and steady manner for intermediate values of the NPAC.

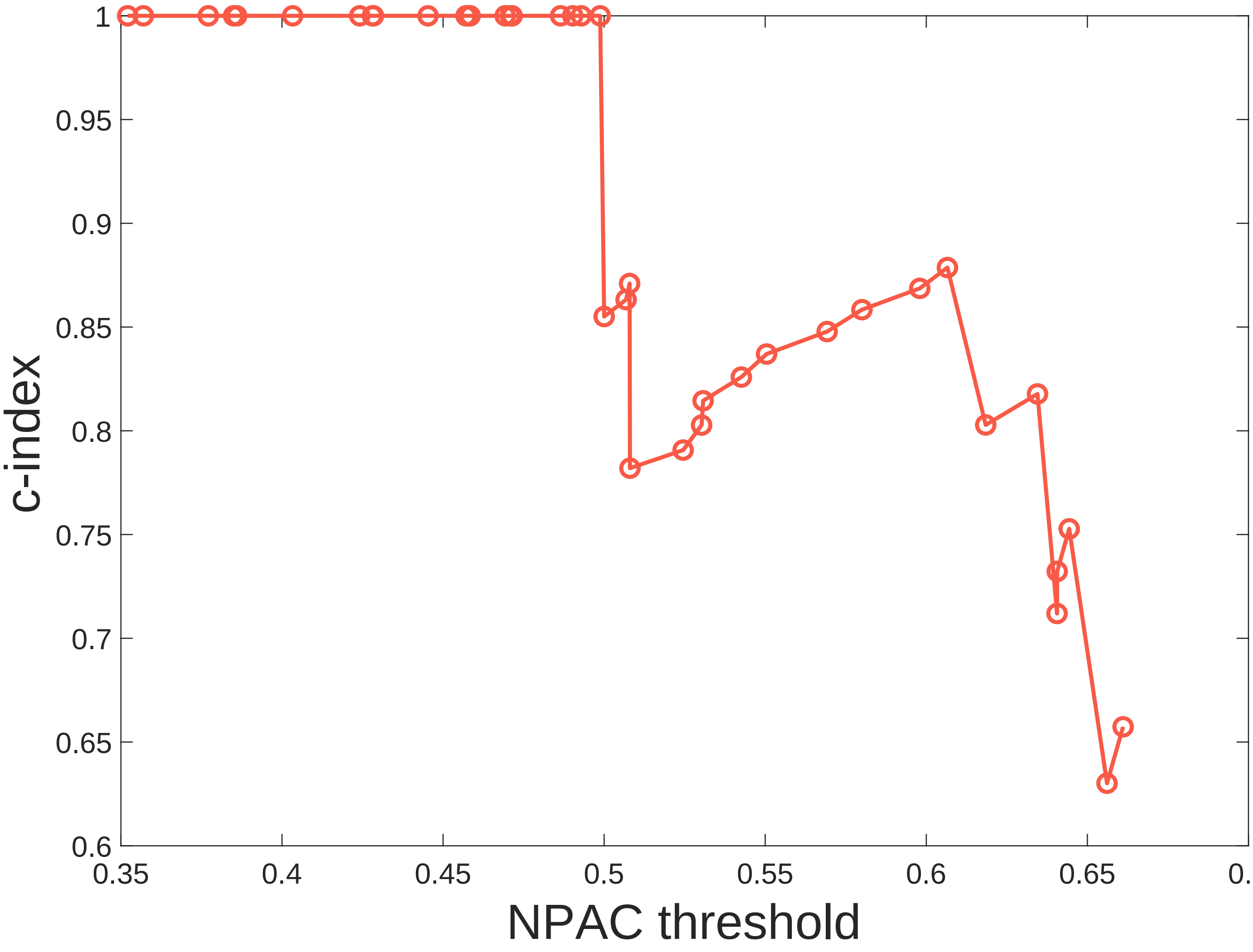


**Fig. S7.** To perform the survival analysis we evaluate all the possible thresholds of NPAC, separating the patients in two different groups according to their NPAC in relation to each threshold. For each of those thresholds, a Kaplan-Meier analysis is performed and the Harrell's c-index is calculated to evaluate the risk associated to patients with dissimilar NPAC. In the figure, the different c-indexes are plotted against the different NPAC thresholds evaluated for the breast cancer dataset. Only groups where the number of patients in the largest one is at most five times bigger than in the smallest one are considered. It is evident that the separation in survival groups by the NPAC is able to distinguish between groups of high risk (high NPAC) and low risk (low NPAC). Steady results with c-index=1 are found meaning that the separation in two groups by its relative position to a NPAC threshold is able to distinguish a subgroup of patients who do not die from the disease and its associate to a low NPAC.

#### 2. *In silico* Survival Analysis

**A. Stochastic mesoscopic model and simulations.** In order to perform simulations where realistic tumor volumes are quickly reached, we developed a hybrid multi-scale stochastic model where whole clonal populations are followed instead of individual cells. The spatial domain was set as a 3D grid discretized in cubic compartments (voxels) of side length $\Delta x$, fixed at 1 mm. This allows to obtain results comparable to available medical images while gathering a sufficiently large number of cells. Four main processes, namely cell division, death, migration and mutation, were implemented as probabilistic events dependent on the local occupation, modeled as a proportion of the carrying capacity $K$ (different in value to that of the previous model) as follows

$$P_{\text{rep}} = \frac{\Delta t}{\tau_{\text{rep}}}\left(1 - \frac{n+d}{K}\right), \tag{1a}$$

$$P_{\text{mig}} = \frac{\Delta t}{\tau_{\text{mig}}\Delta x^2}\left(\frac{n+d}{K}\right), \tag{1b}$$

$$P_{\text{kill}} = \frac{\Delta t}{\tau_{\text{kill}}}\left(\frac{n+d}{K}\right), \tag{1c}$$

$$P_{\text{mut}} = \frac{\Delta t}{\tau_{\text{mut}}}\left(\frac{n_{\text{cpop}}}{K}\right). \tag{1d}$$

where $n$ is the number of alive cells in the population and $d$ denotes the number of total dead tumor cells inside a given voxel. Parameters $\tau$ represent the characteristic times of each process.

Probabilities in Eq. (1) are calculated at each time step and voxel yielding the number of cells undergoing mitosis, apoptosis, mutation and migration for every clonal population. The basic cellular events can be conceptualized as Bernoulli experiments, in the sense that there are two possible outcomes (e.g. to proliferate or not). Following this, the total numbers of proliferating and dying cells from one subpopulation are drawn from binomial distributions $B(n_{\text{cpop}}, P_{\text{rep}})$ and $B(n_{\text{cpop}}, P_{\text{kill}})$. Sample size is equal to the number of cells in that subpopulation $n_{\text{cpop}}$. Number of migrating cells is also drawn from the respective binomial distribution $B(n_{\text{cpop}}, P_{\text{mig}})$ and then they are distributed around a neighborhood of 26 voxels (Moore neighborhood) according to a multinomial distribution. Each accessible voxel is given a probability depending on its distance to the voxel of origin, thus reproducing fickian diffusion. Finally, given that mutations are less frequent events, only one cell at most is allowed to undergo any genotypic transition at a given time step. Mutations allow a cell to move from one clonal population to another, following a mutational landscape that is set up as a Markov chain. Subsequent alterations bring selective advantages by modifying the rate at which cells undergo all processes. The outcome of this set of rules is a scenario in which clonal populations appear, grow and evolve competing for space and resources.

The code was written in Julia (v. 1.1.1), and simulations performed on a network of 12-core 64 GB memory 2.7 GHz Mac Pro and 12-core 128 memory 2.0 GHz iMac Pro machines. Computational cost per simulation was around 5 minutes per run in single-processor mode. Data processing and graphical analysis of simulations were performed in Matlab (R2018a, The MathWorks, Inc., Natick, MA, USA).

**B. *In silico* survival analysis.** With data collected from the 100 breast cancer simulations and 100 lung cancer simulations, a survival analysis was performed, in order to evaluate how the model is able to reproduce PAC and NPAC dynamics and prognostic value observed in reality. Note that this analysis is not a true survival analysis, as it does not rely on real patients. To perform a survival analysis over simulation data, a survival time had to be assigned to each run, as *in silico* tumors are not associated with patients. Survival time depended on diagnostic time, which was randomly selected from ranges extracted from available literature (0.3-5 $\text{cm}^3$ for breast tumors (1), and 0.2-15 $\text{cm}^3$ for lung tumors (2)); and also on death time, which was determined as the time at which simulations reached a certain volume, considered to be incompatible with life (50 $\text{cm}^3$ for breast cancer, and 120 $\text{cm}^3$ for lung cancer (3)). The difference between death and diagnostic time is survival time. For correlation and Kaplan-Meier analysis, prognostic variables were evaluated at diagnostic time. MTV and MSR were calculated by counting the number of active voxels a tumor has. TLG and $\text{SUV}_{max}$ were calculated as the total number of newborn cells and the maximum number of newborn cells found in most active voxel, respectively.

For Kaplan-Meier analysis, a splitting threshold was selected in order to create two different groups of simulated tumors. Survival curves depicted in figures below show the results for the best splitting threshold (splitting threshold with lowest p-value).

***B.1. Breast.***

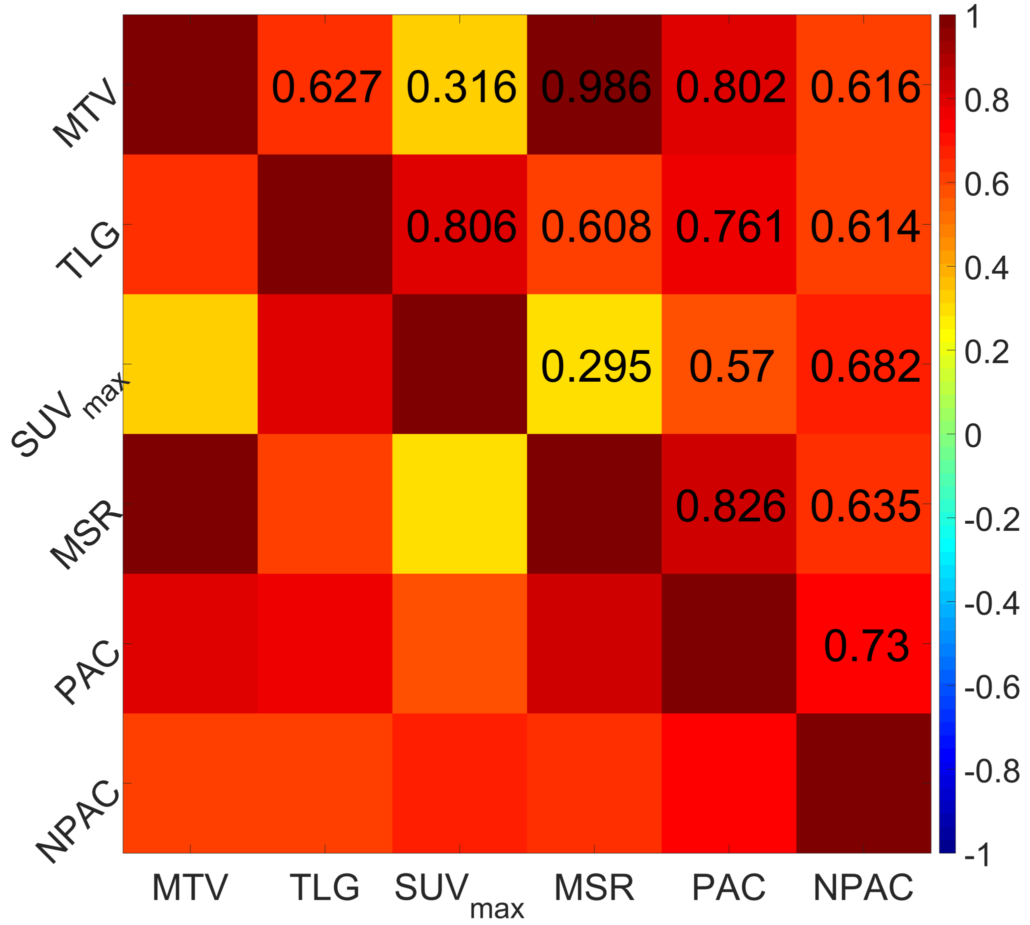


**Fig. S8.** Correlation between main prognostic variables for simulated breast tumors.

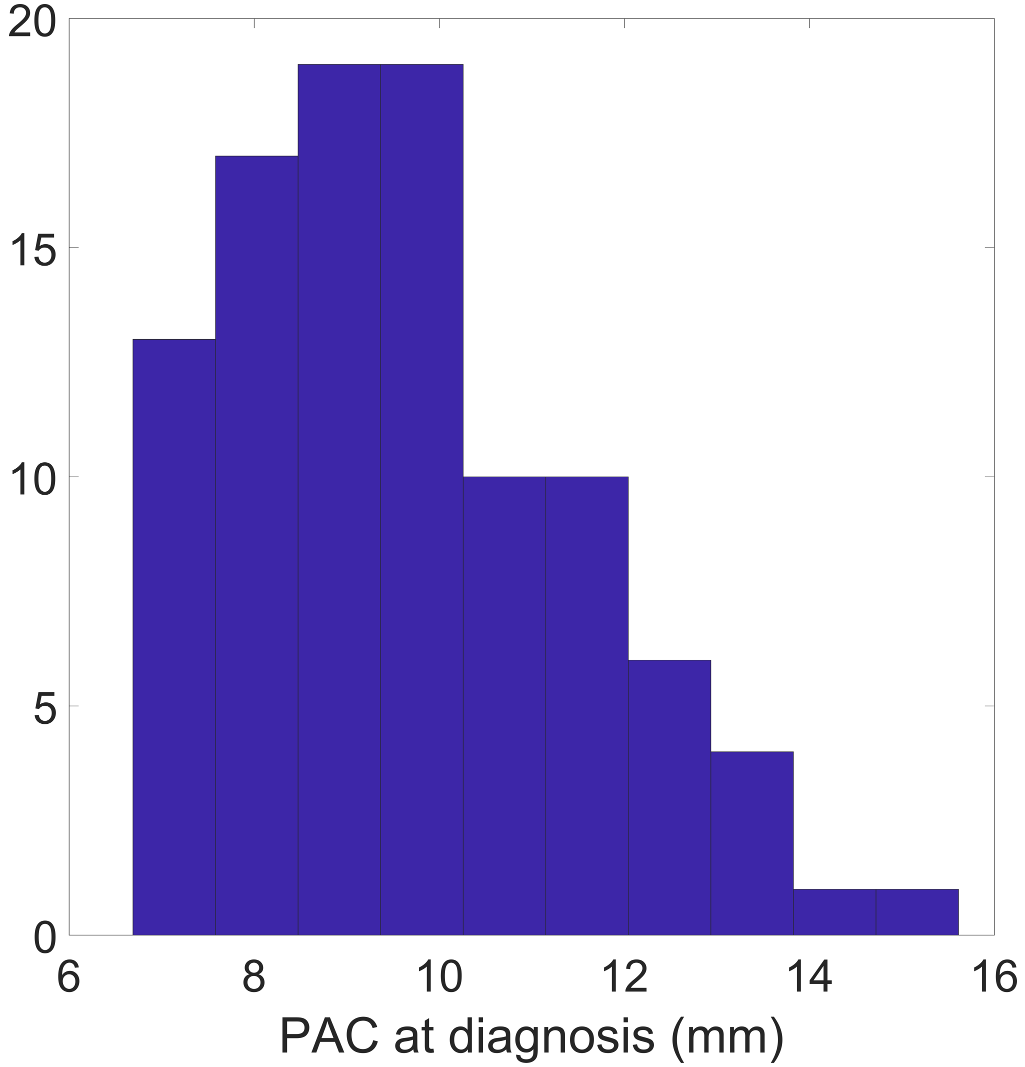


**Fig. S9.** PAC values taken at diagnostic for simulated breast tumors.

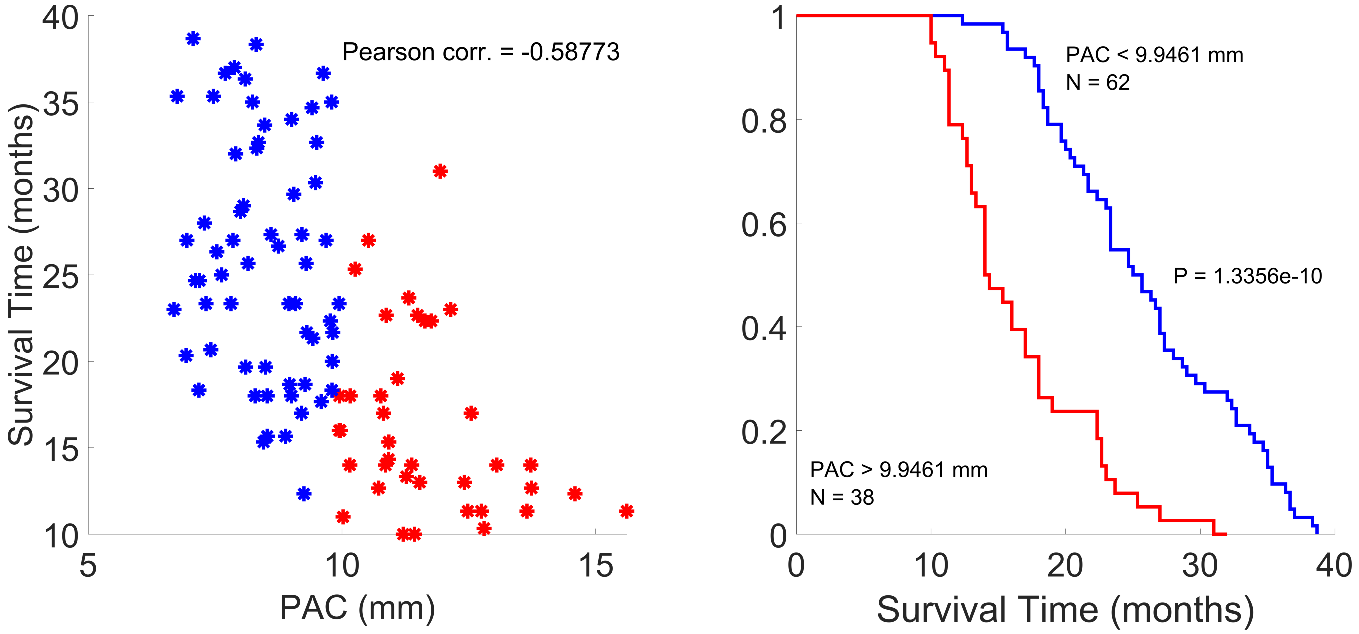


**Fig. S10. Left.** Scatterplot of PAC and survival time for simulated breast tumors. In red, simulated tumors belonging to the group with higher PAC. In blue, simulated tumors belonging to the group with lower PAC. **Right.** Kaplan-Meier analysis of PAC for simulated breast tumors. *In silico* tumors with higher PAC have a lower survival time associated.

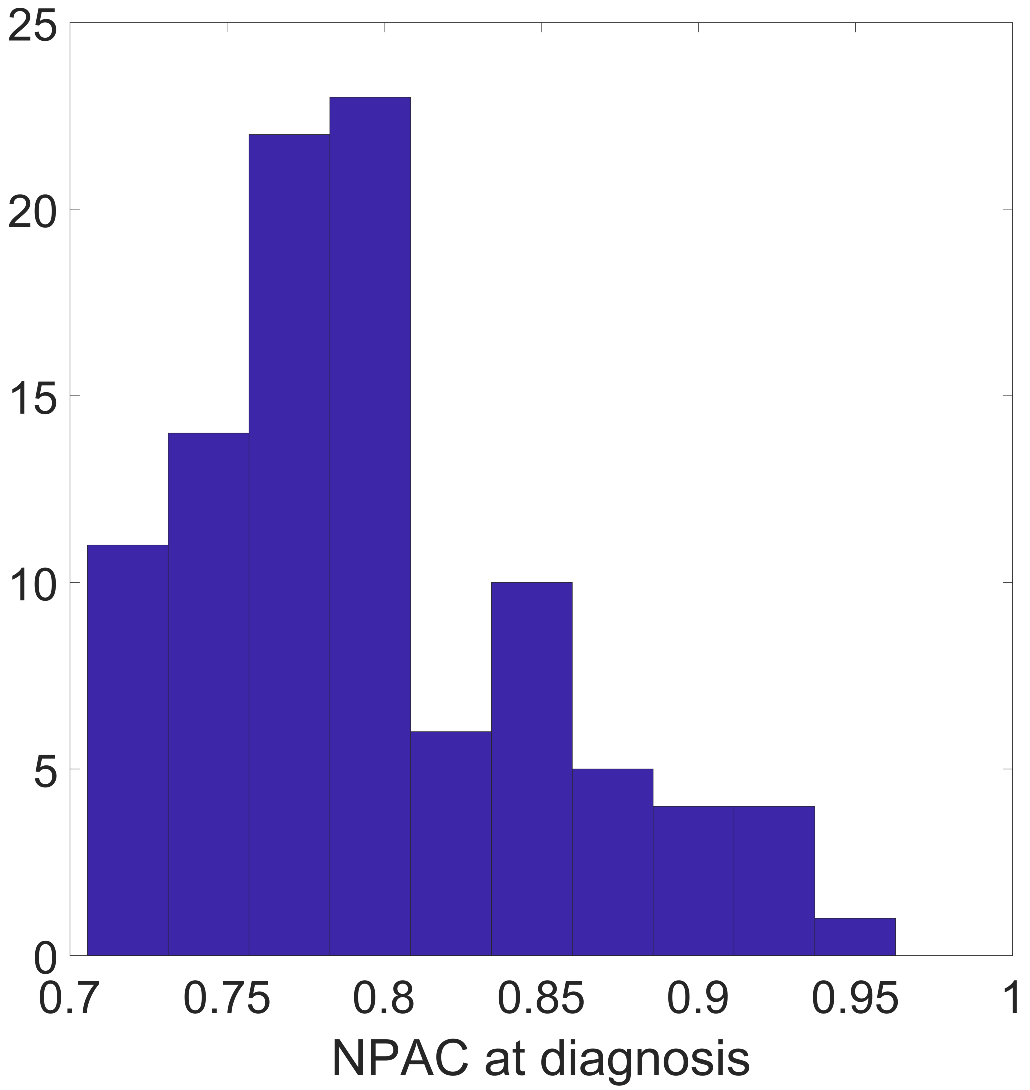


**Fig. S11.** NPAC values taken at diagnostic for simulated breast tumors.

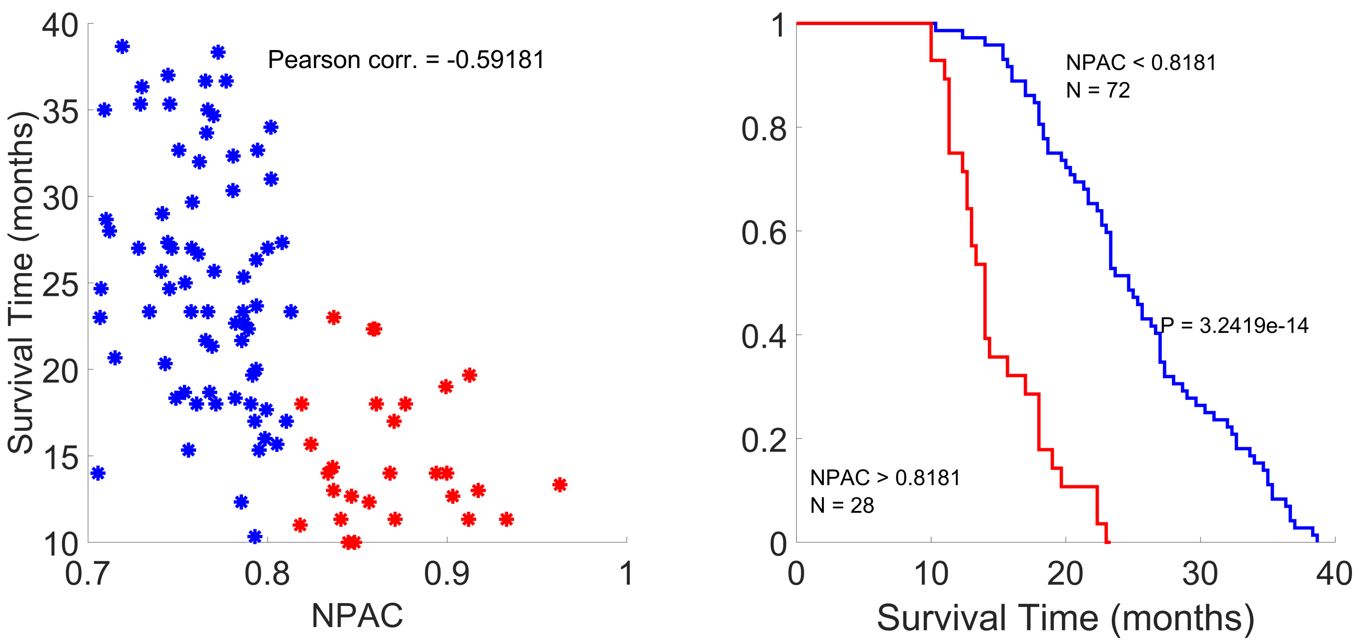


**Fig. S12. Left.** Scatterplot of NPAC and survival time for simulated breast tumors. In red, simulated tumors belonging to the group with higher NPAC. In blue, simulated tumors belonging to the group with lower PAC. **Right.** Kaplan-Meier analysis of NPAC for simulated breast tumors. *In silico* tumors with higher NPAC have a lower survival time associated.

##### *B.2. Lung.*

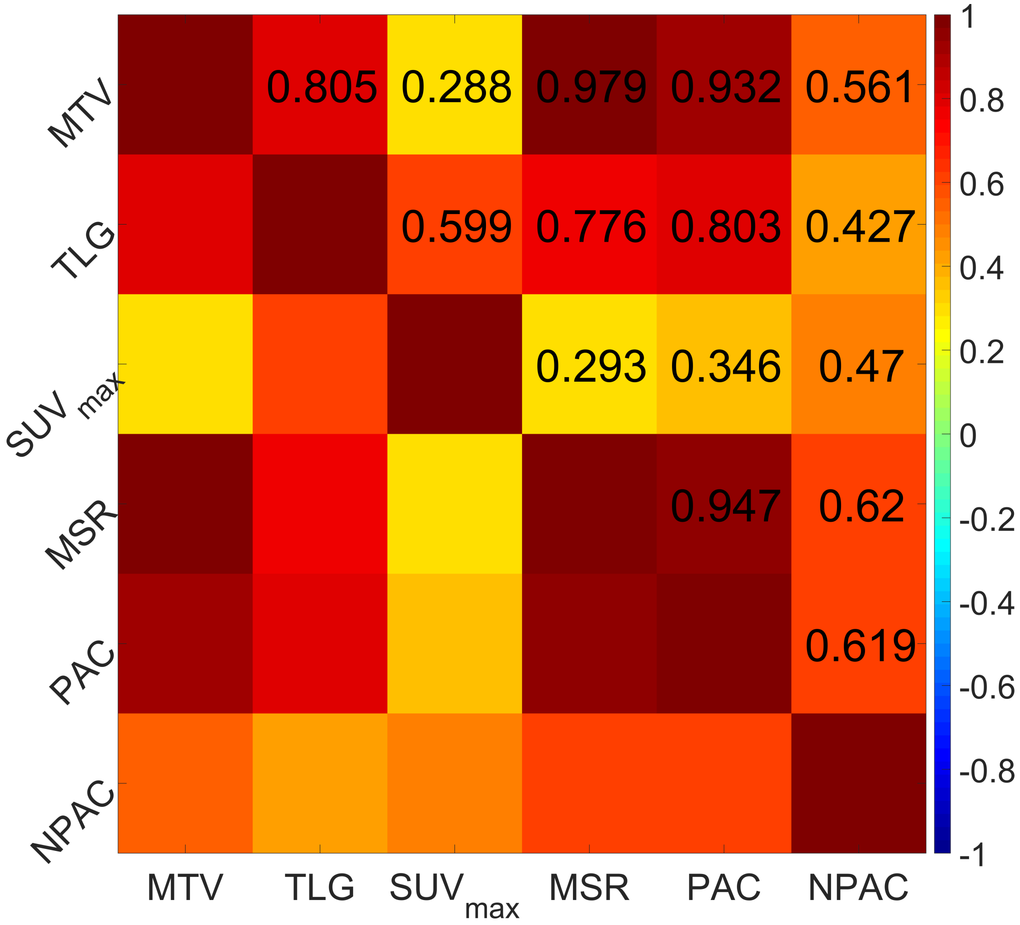


**Fig. S13.** Correlation between main prognostic variables for simulated lung tumors.

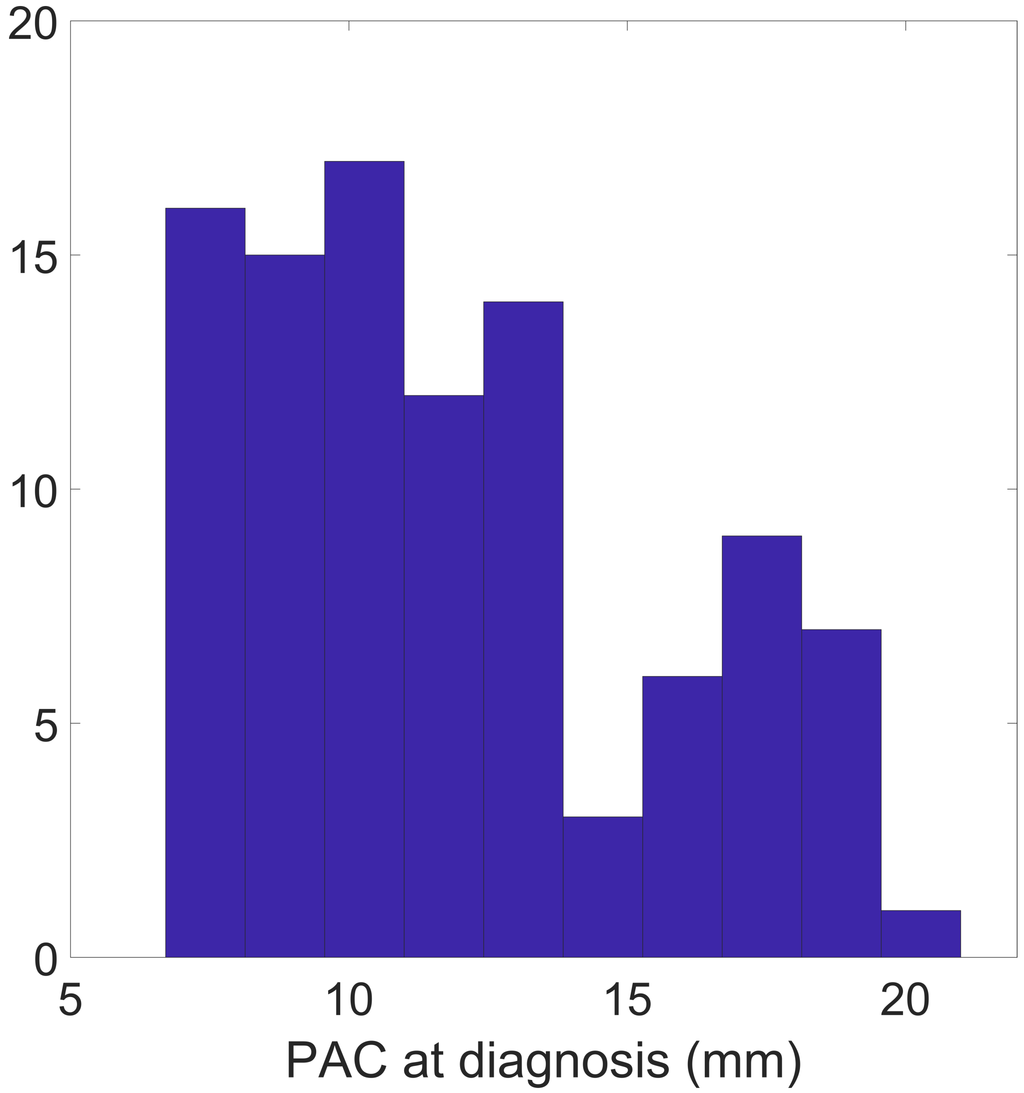


**Fig. S14.** PAC values taken at diagnostic for simulated lung tumors.

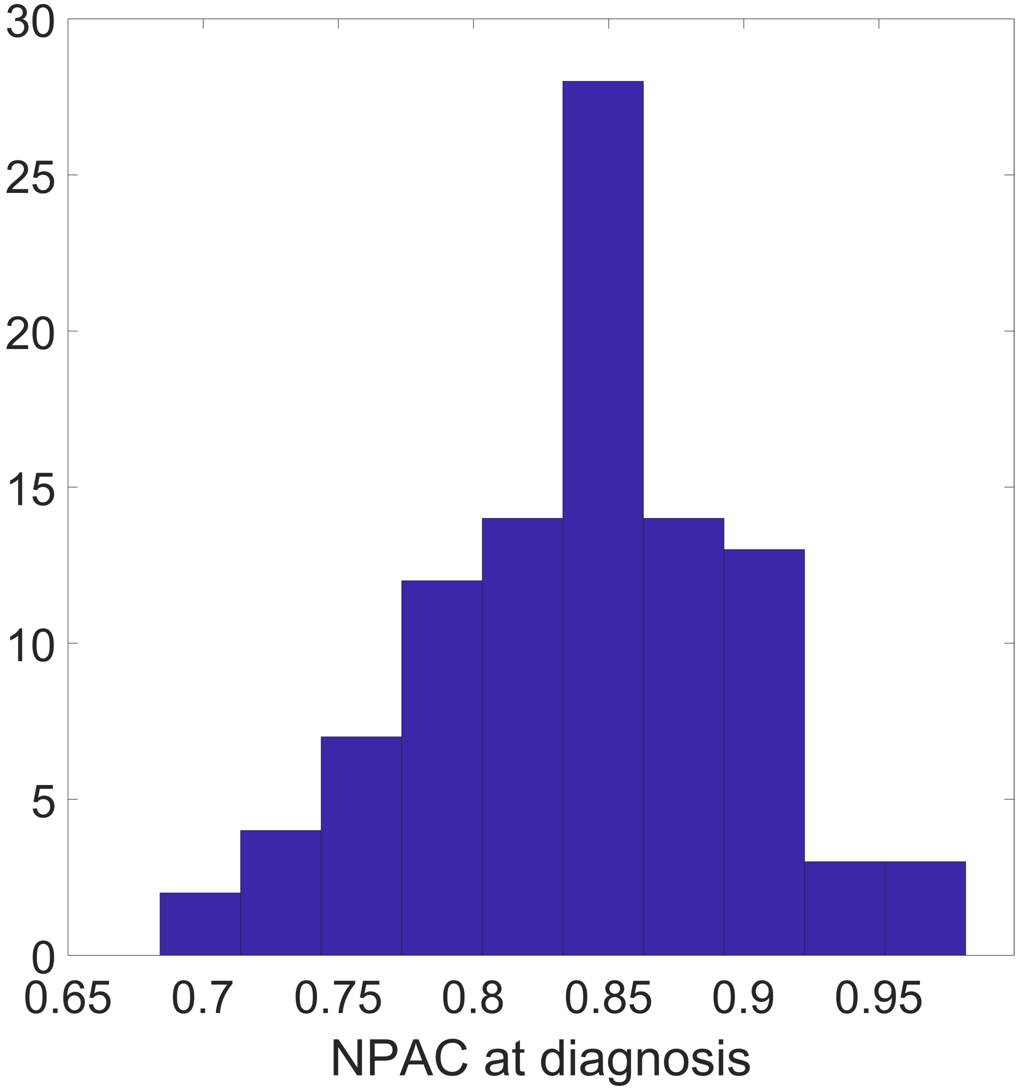


**Fig. S15. Left.** Scatterplot of PAC and survival time for simulated lung tumors. In red, simulated tumors belonging to the group with higher PAC. In blue, simulated tumors belonging to the group with lower PAC. **Right.** Kaplan-Meier analysis of PAC for simulated lung tumors. *In silico* tumors with higher PAC have a lower survival time associated.

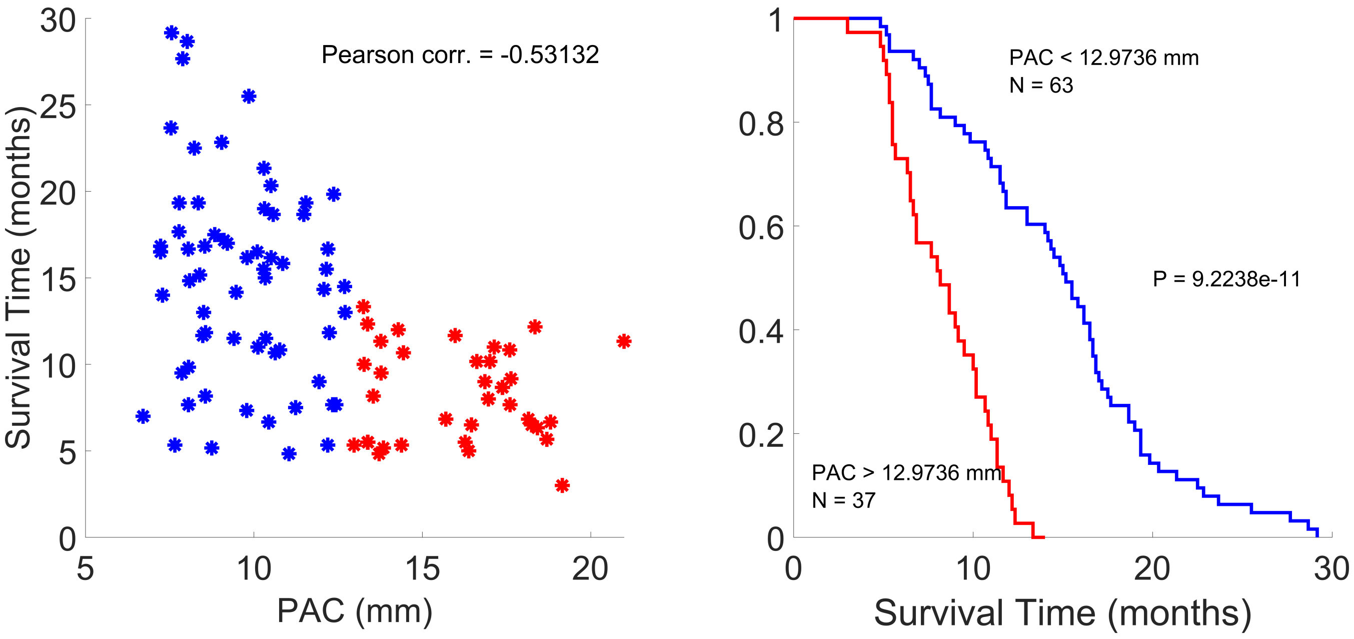


**Fig. S16.** NPAC values taken at diagnostic for simulated lung tumors.

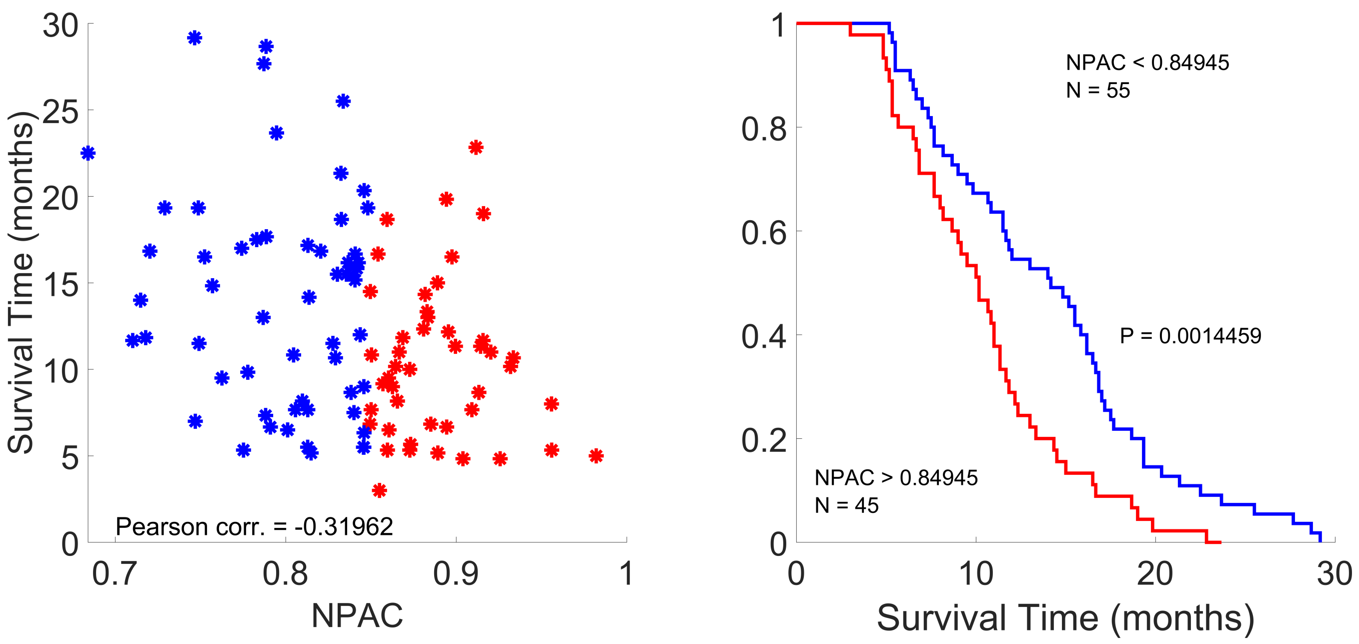


**Fig. S17. Left.** Scatterplot of NPAC and survival time for simulated lung tumors. In red, simulated tumors belonging to the group with higher NPAC. In blue, simulated tumors belonging to the group with lower PAC. **Right.** Kaplan-Meier analysis of NPAC for simulated lung tumors. *In silico* tumors with higher NPAC have a lower survival time associated.

**Movie S1. Type legend for the movie here.**

**Movie S2. Type legend for the other movie here. Adding longer text to show what happens, to decide on alignment and/or indentations.**

**Movie S3. A third movie, just for kicks.**

**SI Dataset S1 (dataset_one.txt)**

Type or paste legend here.

**SI Dataset S2 (dataset_two.txt)**

Type or paste legend here. Adding longer text to show what happens, to decide on alignment and/or indentations for multi-line or paragraph captions.
